# Forecasting intensive care unit demand during the COVID-19 pandemic: A spatial age-structured microsimulation model

**DOI:** 10.1101/2020.12.23.20248761

**Authors:** Sebastian Klüsener, Ralf Schneider, Matthias Rosenbaum-Feldbrügge, Christian Dudel, Elke Loichinger, Nikola Sander, Andreas Backhaus, Emanuele Del Fava, Janina Esins, Martina Fischer, Linus Grabenhenrich, Pavel Grigoriev, André Grow, Jason Hilton, Bastian Koller, Mikko Myrskylä, Francesco Scalone, Martin Wolkewitz, Emilio Zagheni, Michael M. Resch

## Abstract

**Background:** The COVID-19 pandemic poses the risk of overburdening health care systems, and in particular intensive care units (ICUs). Non-pharmaceutical interventions (NPIs), ranging from wearing masks to (partial) lockdowns have been implemented as mitigation measures around the globe. However, especially severe NPIs are used with great caution due to their negative effects on the economy, social life and mental well-being. Thus, understanding the impact of the pandemic on ICU demand under alternative scenarios reflecting different levels of NPIs is vital for political decision-making on NPIs.

**Objective:** The aim is to support political decision-making by forecasting COVID-19-related ICU demand under alternative scenarios of COVID-19 progression reflecting different levels of NPIs. Substantial sub-national variation in COVID-19-related ICU demand requires a spatially disaggregated approach. This should not only take sub-national variation in ICU-relevant disease dynamics into account, but also variation in the population at risk including COVID-19-relevant risk characteristics (e.g. age), and factors mitigating the pandemic. The forecast provides indications for policy makers and health care stakeholders as to whether mitigation measures have to be maintained or even strengthened to prevent ICU demand from exceeding supply, or whether there is leeway to relax them.

**Methods:** We implement a spatial age-structured microsimulation model of the COVID-19 pandemic by extending the Susceptible-Exposed-Infectious-Recovered (SEIR) framework. The model accounts for regional variation in population age structure and in spatial diffusion pathways. In a first step, we calibrate the model by applying a genetic optimization algorithm against hospital data on ICU patients with COVID-19. In a second step, we forecast COVID-19-related ICU demand under alternative scenarios of COVID 19 progression reflecting different levels of NPIs. We apply the model to Germany and provide state-level forecasts over a 2-month period, which can be updated daily based on latest data on the progression of the pandemic.

**Results:** To illustrate the merits of our model, we present here “forecasts” of ICU demand for different stages of the pandemic during 2020. Our forecasts for a quiet summer phase with low infection rates identified quite some variation in potential for relaxing NPIs across the federal states. By contrast, our forecasts during a phase of quickly rising infection numbers in autumn (second wave) suggested that all federal states should implement additional NPIs. However, the identified needs for additional NPIs varied again across federal states. In addition, our model suggests that during large infection waves ICU demand would quickly exceed supply, if there were no NPIs in place to contain the virus.

**Conclusion:** Our results provide evidence for substantial spatial variation in (1) the effect of the pandemic on ICU demand, and (2) the potential and need for NPI adjustments at different stages of the pandemic. Forecasts with our spatial age-structured microsimulation model allow to take this spatial variation into account. The model is programmed in R and can be applied to other countries, provided that reliable data on the number of ICU patients infected with COVID-19 are available at sub-national level.

## Introduction

The COVID-19 pandemic has a profound impact on health care demand. The high communicability of the virus and the substantial share of severe cases pose the imminent risk of health care systems being overwhelmed, thus putting the safe and adequate provision of health care at risk (Ferguson et al. 2020). A shortage of beds and staff is particularly relevant as it pertains to beds in intensive care units (ICUs), given that COVID-19 patients with severe respiratory symptoms frequently require intubation and mechanical ventilation (Rodriguez-Morales et al. 2020; Stang et al. 2020a; McCabe et al. 2020). Mitigation policies have been put in place that aim at suppressing infections and thus reducing peak health care demand. As long as COVID-19 vaccination is not widely available, non-pharmaceutical interventions (NPIs) will play a key role in containing the pandemic (Han et al. 2020). NPIs include, for example, isolation of symptomatic cases, physical distancing, wearing face masks, school closures, as well as stay-at-home orders (e.g. lockdowns) (Chowdhury et al. 2020; Haug et al. 2020).

While it is difficult to isolate effects of singular NPIs on the pandemic, there is by now ample evidence that NPIs are generally an adequate strategy to reduce the reproduction number *R*_*t*_, and thus the daily numbers of new infections, hospitalizations, and deaths related to COVID-19 (Flaxman et al. 2020; Li et al. 2020a; Dehning et al. 2020). Whereas NPIs are effective in reducing the transmission of COVID-19, interventions such as stay-at-home orders and school closures can have profound negative effects on the economy, social life and individual well-being (Fuchs-Schündeln, Kuhn, & Tertilt 2020; Naumann et al. 2020; Bäuerle et al. 2020; Bauer & Weber 2020). Therefore, policy makers and other stakeholders aim to use NPIs with caution, while reducing the spread of COVID-19 sufficiently in order not to overburden health care systems.

Epidemiological models that forecast the spread of COVID-19 have been used in several countries to inform policymaking on the efficacy of past and future policy measures (see, for example, Alban et al. 2020; Davies et al. 2020; IHME et al. 2020; IHME & Murray 2020; Jewell et al. 2020; Kucharski et al. 2020; Nadler et al. 2020; Prem et al. 2020; Ruktanonchai et al. 2020; McCabe et al. 2020, Hellewell et al. 2020). Forecasts of infectious diseases are often based on compartmental mathematical models, which divide a population into states and assume rates of transition from one state to another (Brauer 2017). The Susceptible-Exposed-Infectious-Recovered (SEIR) framework that is frequently applied to virus infections uses four states (Susceptible, Exposed, Infectious, and Recovered) to model the spread of the virus. The number of infections and deaths in a population are forecasted as a function of time, and are based on assumptions about the probability of disease transmission, incubation period, recovery rate, fatality rate and so on (see for example Carcione et al 2020; Teslya et al. 2020).

Results of epidemiological models indicate that NPIs can be helpful to avoid a rapid increase in COVID-19 infections and, consequently, rising demand for ICU beds (e.g. Ferguson et al. 2020; Dehning et al. 2020; Römmele et al. 2020). However, it is challenging to determine whether mitigation measures have to be maintained or even strengthened in order to prevent ICU demand from exceeding supply, or whether there is leeway to relax the measures. This challenge, which calls for an extension of conventional epidemiological models, arises for two main reasons: First, conventional epidemiological models often tend not to fully capture the dynamic spread of the virus across space and time (Kapitsinis 2020; Mense & Michelsen 2020; Thomas et al. 2020). However, capturing this dynamic is crucial given the high spatial and temporal variation of the COVID-19 pandemic that often results in sub-national infection hot spots. The spatial clustering of severe cases can put significant strain on health care in these areas (Thomas et al. 2020). In order to adequately forecast spatial variation in ICU demand, it is also not sufficient just to account for the observed spatial variation. Whether specific places are likely to see rises in ICU demand in the near future is not only dependent on the level of infections in these places, but also to some degree dependent on the level of infections in places with which these places have high social interaction. Thus, it is important to account for likely spatial diffusion pathways of the pandemic in the forecasts. In addition, spatial variation in the impact of COVID-19 on ICU bed demand is also dependent on the sub-nationally varying age structure of the population. The higher the median age of a population, the higher the impact tends to be in terms of severe cases and thus the burden on health care facilities (Levin et al. 2020). Capturing these aspects requires a spatially disaggregated modelling approach, which also takes into account that NPIs vary across space and time. The second reason why it is challenging to study the effects of NPIs on ICU demand is that temporal variation in ICU demand not only depends on the level of NPIs. It also depends on a series of other factors that include, but are not limited to, seasonality, advances in medical prevention and treatment, as well as the age-specific pattern of infections (see also Haug et al. 2020).

The main aim of this paper is to cope with these challenges by developing and applying a spatially disaggregated microsimulation model based on the SEIR framework to forecast future ICU demand over a period of 2 months. The length of the forecasting period is motivated by the fact that the high dynamic of the pandemic with frequent NPI-related government interventions would make it difficult to forecast potential paths over longer periods. Key features of our approach include that we are able to account for spatial variation in population age structure, and for spatial and temporal variation in the COVID-19 dynamic at high geographic detail, including possible spatial diffusion pathways. Related to the effects of NPIs, we decided not to model the link between NPIs and ICU demand directly, as it would be very challenging to separate NPI effects from other factors mentioned above (e.g. seasonality effects, etc.). Instead, our model attempts to estimate the total effect of all factors that influence the reproduction number, and how this total effect varies across space and time. This then allows us to derive forecasts based on different scenarios that can be mitigated through strengthening or weakening NPIs. If, e.g., we estimate for a region a reduction of the reproduction number by 40% in the last period before the forecast, we can keep this effect for the forecast constant, or reduce or increase it under the assumption that NPIs are adjusted in the forecast period.

The microsimulation specification provides more freedom in accounting for complexity in spatial diffusion processes compared to frequently used differential equation specifications of compartmental models. As such, our spatial microsimulation model complements the conventional compartmental mathematical modelling approach in order to support decision making on the level of NPIs required at the national and sub-national level at different phases of the pandemic. Overall, our goal is to propose a model that captures the spatial and temporal variability of COVID-19 infections and health care demand, while being sufficiently intuitive to inform policy makers and health care stakeholders.

We calibrate our model against the daily total number of ICU patients and then forecast the number of ICU patients over a period of 2 months. ICU data is a more reliable indicator for assessing healthcare demand than incidence data as one big limitation of incidence data is that they tend to undercount new infections due to a considerable number of asymptomatic and mild cases and a lack of systematic testing (Ioannidis, Cripps and Tanner 2020; Chin et al. 2020, Moghadas et al. 2020; Li et al. 2020b; Wu and McGoogan 2020; Holmdahl and Buckee 2020, Bokh-Ewald et al. 2020, Streeck et al., 2020). We therefore apply our model to data from the novel German intensive care register, which was established in March 2020 by the German Interdisciplinary Association for Intensive Care and Emergency Medicine (DIVI) and the Robert Koch-Institute (RKI 2020a; RKI 2020b). The simulation model can be applied to other countries where data on the number of ICU patients with COVID-19 are available.

Using Germany as a case study, we demonstrate that our model captures the impact of the COVID-19 pandemic on health care demand in the context of NPIs and other influencing factors. Our forecasts for a quiet summer phase with low infection rates identified quite some variation in potential for relaxing NPIs across the federal states. By contrast, our forecasts during a phase of quickly rising infection numbers in autumn (second wave) suggested that all federal states should implement additional NPIs. However, the identified needs for additional NPIs varied again across federal states. In addition, our model suggests that during large infection waves ICU demand would quickly exceed supply, if there were no NPIs in place to contain the virus.

## Background

To make our results more understandable for an international readership, we provide some background information on the course of the COVID-19 pandemic in Germany. The number of infections started to rise steeply for the first time in March 2020, and particularly southern and western states were affected during the first COVID-19 wave. The nationwide number of ICU patients with COVID-19 during the first wave peaked at around 3,000 individuals in the beginning of April, and decreased substantially to around 200 individuals during the summer months. A resurgence in infections beginning in September resulted in constantly increasing numbers of ICU patients. In this wave, next to the states already heavily affected in the first wave, also the southern part of eastern Germany witnessed stark rises in incidence rates. By the middle of December, more than 5,000 ICU patients with COVID-19 were reported nationwide.

Compared to many neighboring countries Germany suffered relatively few COVID-19 deaths per 100,000 inhabitants between March and December 2020 (Stang et al. 2020b). This can be traced to a number of factors: the relatively high availability of COVID-19 tests, the relatively low median age of the early cases, the introduction of severe NPIs already in March, and the fact that Germany has the highest number of ICU beds per 100,000 people in Europe (Rhodes et al. 2012). Across the country, schools and childcare facilities closed mid-March; and while social distancing was not yet mandatory, it was strongly recommended. On March 23, a far-reaching nationwide contact ban was imposed by government authorities, which included the prohibition of small public gatherings as well as the closing of restaurants and all nonessential stores. Facemasks became mandatory for the use in shops and public transport by the end of April (Dehning et al. 2020; Naumann et al. 2020). The number of infections slowed down in the second half of April, so that some measures were relaxed in mid-May. Schools and restaurants reopened in late spring. Apart from some local resurgences, infection rates remained rather low during summer. However, the beginning of a second wave of infections hit Germany in late September. To reduce the spread of the virus, a partial nationwide lockdown was imposed on November 2, including the closure of restaurants, entertainment facilities, and public recreation centers. In addition, gatherings in public places were limited to members of two households. On December 16, the lockdown was extended including the closure of schools, childcare facilities, and nonessential stores.

Germany is organized politically as a federal republic consisting of sixteen partly sovereign federal states representing the German NUTS-1 regions, with the responsibility for implementing NPI policies mostly lying with the federal state governments. The federal sovereignty caused some sub-national variation in NPIs during the summer months. This was primarily due to states with lower infection levels imposing fewer NPIs than states with higher infection rates. In part, also the level of the 401 districts representing the German NUTS-3 regions can play a role in containment policies: If one of the 401 districts in Germany exceeds a certain incidence over 7 days per 100.000 population, additional lockdown measures may be applied in this district.

## Modelling Approach

### Overview

We use a spatial age-structured microsimulation model implemented in the R software environment to forecast ICU demand under alternative scenarios of the future dynamic of COVID-19. The time unit of the model are days. At the core of our extended model is a microsimulation based on the SEIR framework, in which individuals can obtain seven distinct states (see e.g. an der Heiden & Buchholz 2020). (1) Susceptible individuals can become (2) latently infected through contact with infectious individuals. After this period of latency, the infected individuals enter a state in which they are (3) infectious but not yet considered ill, which is followed by an (4) infectious state of symptomatic or asymptomatic illness. From this state, individuals can either move to the states (5) dead, (6) admitted to ICU, or (7) recovered. Individuals in state (6) admitted to ICU, can either move to the states (5) dead or (7) recovered. The simulation states and transitions are depicted in Figure 1. A detailed description of the equations and the parameters that define state durations and age-specific transition probabilities between these states is provided in Appendix 1 and Table A1 together with the corresponding references.

**Figure 1:**
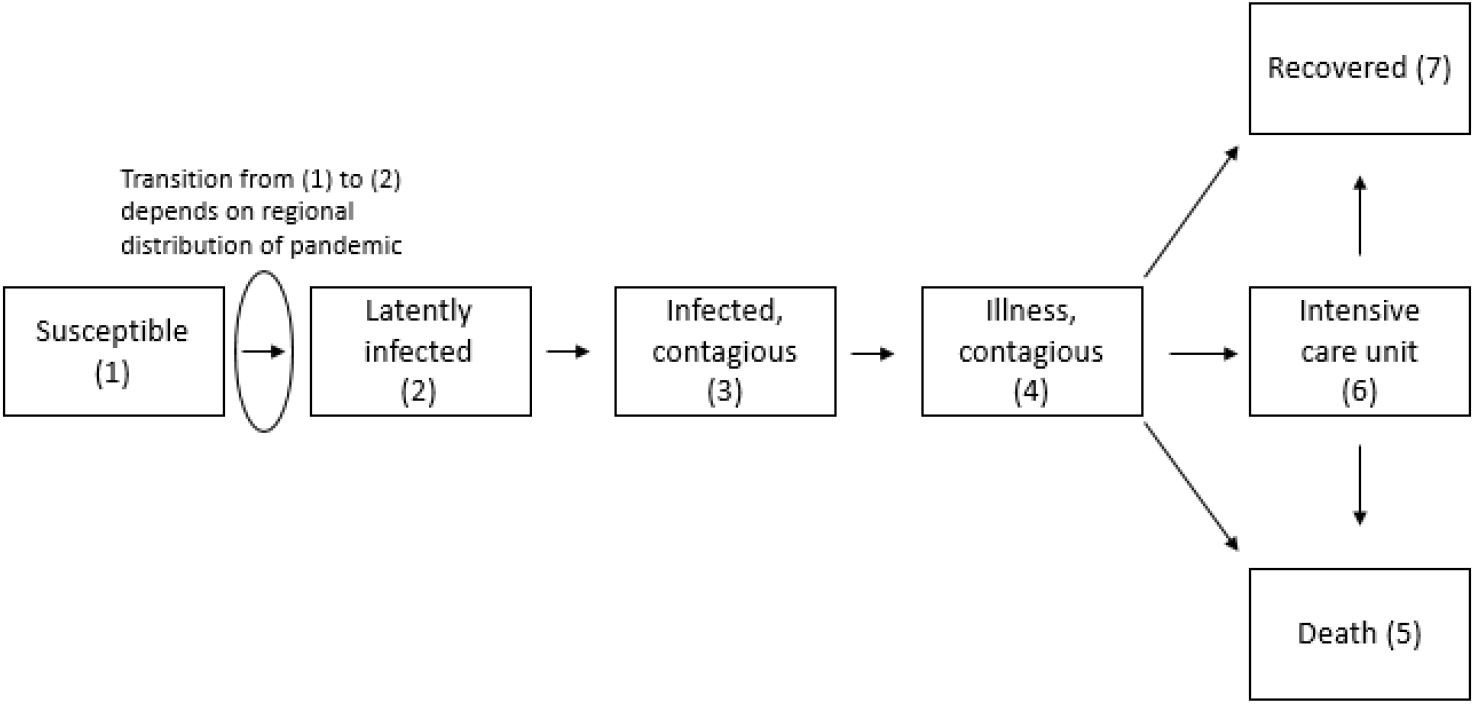
Simulation states and transitions.

We contribute to the existing literature by proposing and implementing a microsimulation model that extends conventional compartment models of COVID-19 through the addition of three interrelated aspects. First, while many conventional models are run at the national level, ICU demand assessments require a more spatially refined approach (Thomas et al. 2020). We therefore run our model at a high level of geographic detail at the level of the 401 districts, which is equal to the standardized NUTS-3 level hierarchy of regions of the European Union (see also Figure 1A in Appendix 1). This allows us to account for spatio-temporal variation in the dynamic of the pandemic and spatial variation in population age structures. To derive our representative population for the 401 districts by 5-year age groups, we use data from the Federal Statistical Office of Germany. In this paper, the simulations are based on a 20% sample of the German population (16,603,842) which we found to be a good compromise between undisturbed infection dynamics and computational effort. Second, for more realistic spatially-refined forecasts, we do not only take the current spatial distribution of the pandemic into account, but also potential spatial diffusion pathways. For that reason, we specify infections risks (i.e., transition from (1) susceptible to (2) latently infected) in such a way, that they are not only dependent on the share of contagious infected persons in the district *i* of residence, but to some degree also on the share of contagious infected persons in districts *j* with which district *i* has high social connectivity. As proxy for social connectivity we use district-level commuter flow data derived from the German Federal Employment Agency (2019). Such data have been shown to strongly correlate with mobile phone activity data (Tizzoni et al. 2014). Third, while many compartmental models rely on incidence data, we calibrate our model against trends in ICU patients with COVID-19 (RKI 2020a) as these data are more reliable for forecasting ICU demand (Holmdahl and Buckee 2020).

Although we run our model at the level of the 401 districts, we do not implement the calibration at the district level. The main reason is that the ICU occupancy at the district level is not necessarily an indicator of the disease dynamic in this district. Many districts have small or even no ICU capacities, so that patients living in these districts have frequently to be diverted to neighboring districts. Thus, district-level variation in ICU occupancy is rather driven by spatial variation in ICU supply than by spatial variation in ICU demand. For that reason, we decided to calibrate our model at a higher level of spatial aggregation, by contrasting observed and simulated numbers of ICU patients at the level of the 38 NUTS-2 regions of Germany. Sensitivity checks suggest that also at this more coarse level of spatial aggregation we capture variation in the spatial dynamic very well. This decision implies that we estimate spatio-temporal variation in NPIs and other factors that influence the course of the pandemic at this level. Working with 38 regions also has the advantage that we need to estimate fewer parameters in the calibration process. In a first step, we perform the calibration of our model for the observed period before we implement the forecast for different scenarios in a second step. In the present paper, we aggregate the simulation outcomes to the level of the 16 federal states (i.e., NUTS-1, see also Figure A1 in Appendix 1), given that most NPI policies are implemented at this administrative level. Detailed results for the 38 NUTS-2 regions are presented in Appendix 2.

### Parameter and model specification

The parameters that we determine through calibration are denoted with Greek letters, while parameters that we derive from the literature or through sensitivity checks are denoted in Latin letters. We use annual data on district-level commuter flows in 2019 to approximate the spatial diffusion of COVID-19 across regions due to human mobility. The data capture the number of commuters across district boundaries through a symmetric matrix denoting the total gross flows of persons commuting between two districts in each off-diagonal matrix element. In addition, the total number of employees whose workplace and place of residence are located within the same district are added on the matrix diagonal. Second, we derive row-wise proportional weights summing up to 1 in order to capture the degree to which the risk of infection of an individual living in district *i* is not only determined by the level of infections in district *i*, but also by the level of infections in other districts *j* with high social connectivity. The weighting parameter *w_int* moderates to which degree this commuter information is taken into account in our model. A value of 0 implies that full weight is given to the commuter flow matrix described above. A value of 1 implies that the infection risk in district *i* only depends on the level of infections in that specific district (more details are provided in Appendix 1). Sensitivity checks suggest that a *w_int*-value of 0.9 is well suited to predict spatial diffusion patterns in ICU demand reasonably well. It is possible to replace the commuter data with other mobility data reflecting the interconnectivity of districts or similarly disaggregated spatial units (cell phone data, etc.), if such data are available.

Related to the impact of NPIs on ICU demand, we decided for reasons provided in the introduction not to directly model the link between NPIs and ICU demand. Instead, we decided to subsume all factors that have an ICU-demand relevant impact on the basic reproduction number under a single parameter for each region-time combination.

Accordingly, the time-varying reproduction number is calculated as follows:

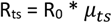

where R_0_ denotes the basic reproduction number. In Germany, R_0_ varies according to estimates of the RKI (2020c) between 3.3 and 3.8, and we use in our models a time-constant value of 3.5. The parameter *μ*_*ts*_ alters the basic reproduction number R_0_ and is calibrated so that our model results for region *s* in week *t* reproduce the observed ICU cases. The parameter *μ*_*ts*_ is the only parameter in our model which is calibrated by an optimization. For reasons discussed above and detailed in Appendix 1, the values of the *μ*_*ts*_ parameter are obtained by calibrating against hospital data on the daily number of ICU patients with COVID-19 at the level of the 38 NUTS-2 regions. Accordingly, the *μ*_*ts*_ parameter does not aim to capture the overall disease dynamic, but the disease dynamic relevant for ICU demand. The objective function for the applied genetic optimization algorithm (Scrucca 2013) is the root mean square of the distances between the daily reported and simulated patients in ICUs. In order to limit the degrees of freedom in our model, we decided to derive all other parameters (see Appendix 1) not based on optimization but on the existing literature or sensitivity analysis. During the first wave in spring 2020, it has been estimated that strict NPIs imposed in most Western European countries reduced pre-intervention values of the time-varying reproduction number by up to 80% (Flaxman et al. 2020; Dehning et al. 2020). Accordingly, we believe that *μ*_*ts*_ represents the impact of NPIs to a substantial but not complete degree.

As the calibration accounts for the disease dynamic only at the level of the 38 NUTS-2 regions, an optional weighting parameter *w_obs* offers the possibility to readjust for district-level variation in the disease dynamic. In this way, it is possible to account for the emergence and waning of hot spots over the course of the pandemic, which enables us to ensure that we enter the forecasting period with a quite realistic spatial distribution pattern at the district level. Having a realistic spatial distribution pattern is an important prerequisite, if we want to take in the forecast potential spatial diffusion pathways adequately into account (through *w_int*). This readjustment, however, is based on incidence data and the assumption that the incidence data provides reliable information on the spatial variation of infection levels; an assumption that for Germany, with its well-developed testing facilities, is in our eyes justified. In other countries, *w_obs* (and *w_int*) should probably be used with more caution. The parameter *w_obs* allows us for the optimization period to redistribute the simulated infections at time *t* in the districts of state *s* based on district-level proportions of reported incidence data for that time *t* (see Figure A10 in Appendix 2). The parameter can vary between 0 and 1. In the cases presented here, *w_obs* was set to 0.5, which implies that when determining the spatial distribution of new infections across the districts at day *t*, 50% of new infections follow the simulated spatial distribution, and 50% follow the distribution in the observed data of registered new infections. Sensitivity checks show that for the state level outcomes presented in this paper, the effect of *w_obs* is rather limited.

### Demonstrating the analytical assets in different states of the pandemic: three cases

While our simulation model can be used for a large range of analytical purposes, we focus here on highlighting the assets of our model through three illustrative cases. The key question is whether and to what degree there is spatial variation across Germany in the level of NPIs necessary to keep the pandemic at a manageable level at different stages of the COVID-19 pandemic. To make reasonable comparisons across federal states, we present the number of ICU-patients with COVID-19 as the share of the total number of ICU beds available in that specific federal state, which we call ICU capacity in the following. For simplicity, we decided to use the total number of ICU beds registered on the 30^th^ of April when hospitals had prepared for the first wave and reported a total number of 32,691 ICU beds nationwide (RKI 2020a). One has to be aware that between 50% and 65% of these beds are typically occupied by patients without COVID-19 (own calculations based on RKI 2020b), which implies that exceeding an ICU-capacity level of 35% for patients with COVID-19 might result in a serious risk of overburdening the ICU health care system.

We present three cases and we run each case simulation 10 times to reduce the influence of randomness. The figures present the mean outcomes of these 10 simulations. Figure 2 illustrates in which periods of the pandemic the three cases are situated. The first case is a hypothetical baseline scenario at the very beginning of the COVID-19 pandemic in Germany, where we let the pandemic spread with an R_ts_ of 3.5 (*μ*_*ts*_ =1). With this we aim to explore spatial variation in how ICU demand would have developed in an uncontained pandemic. The second and third cases show for two different stages of the pandemic how the model can support national and regional decision-makers in assessing whether mitigation measures have to be maintained or even strengthened to prevent ICU demand from exceeding supply, or whether there is leeway to relax them. The second case (“low dynamic”) deals with a period with relatively low infection rates which Germany experienced during the summer months of 2020. The third case (“high dynamic”) is related to a period of resurgence, which occurred in Germany in autumn 2020. In the second and third cases, we first derive estimates of the weekly R_0_-adjustment parameters *μ*_*ts*_ to make assessments on spatio-temporal dynamics of COVID-19. In a second step, we then use this information to forecast the pandemic for different *μ*_*ts*_ scenarios (that can be influenced by NPIs) over a period of 2 months. In the low dynamic case, we show forecasts for a scenario based on a constant *μ*_*ts*_, and for a scenario based on a 30% increase in *μ*_*ts*_. Contrasting these two scenarios allows us to explore spatial variation in potential for relaxing NPIs. In the high dynamic case, we present forecasts for three scenarios: constant *μ*_*ts*_ and a *μ*_*ts*_ decrease of 20% respectively 40%. This allows us to detect variation in the degree to which different federal states should consider additional mitigation measures to ensure that the COVID-19 pandemic remains at a manageable level.

**Figure 2:**
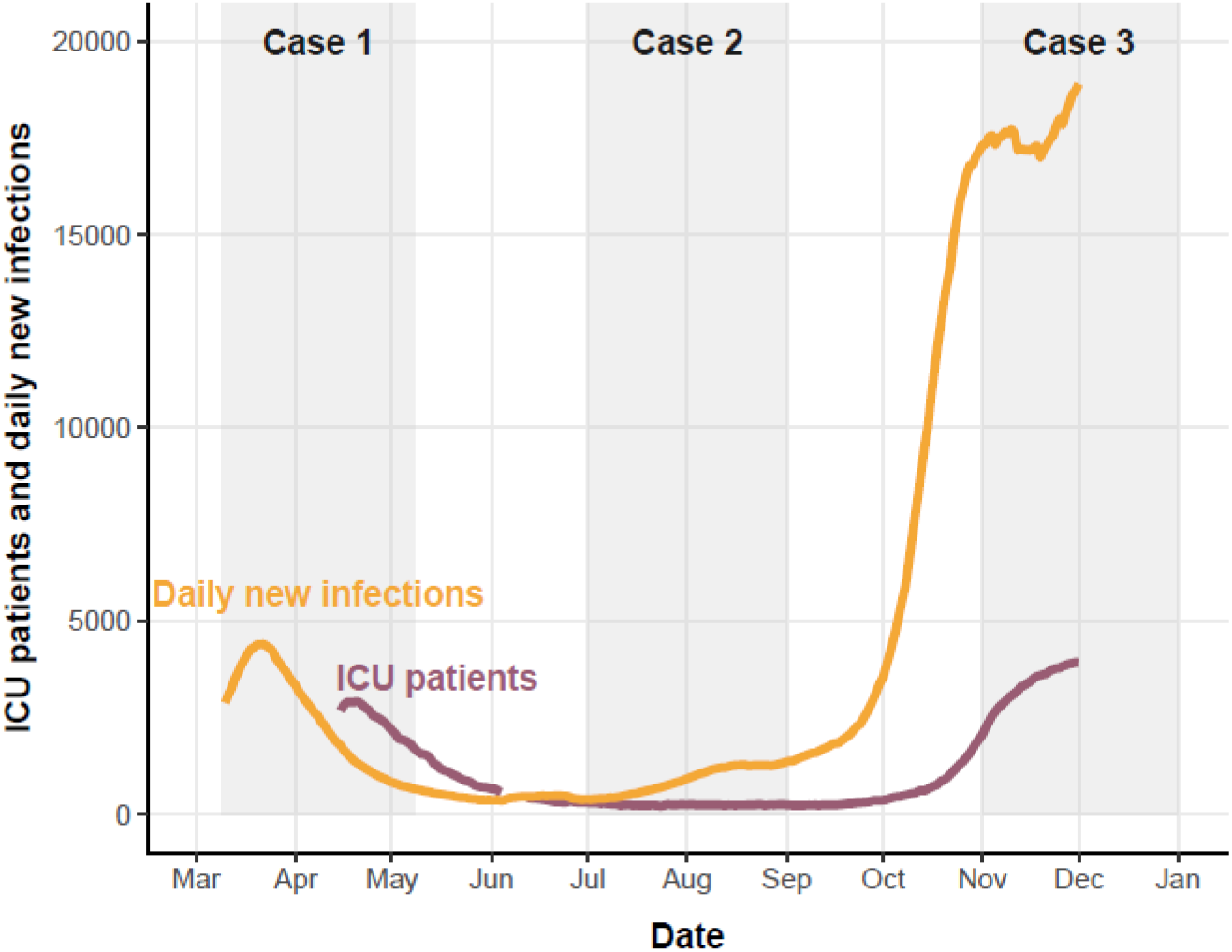
Overview over cases.

## Results

### Case 1: Uncontained pandemic

For the baseline scenario, in which we let the pandemic spread through the country without any restrictions (*μ*_*ts*_ =1; R_ts_ = 3.5), we chose the 9^th^ of March 2020 as the starting date of our simulation. This date marks the beginning of the week when case numbers started to rise drastically in Germany and spurred the implementation of severe NPIs by the end of the week. In addition, at that date all relevant local hot spots of the first wave of the COVID-19 pandemic had already emerged, so that we are able to start with a realistic spatial pattern of our initial conditions. In contrast to the other two scenarios, we do not make use of the ICU statistics for adjusting spatial variation patterns here, as no reliable statistics on ICU-patients in Germany have been available before mid-April. We thus rely exclusively on the recorded infection numbers at the district level in the period around the 9^th^ of March which we use to determine the starting conditions (see Appendix 1 for details). These are likely to be underestimated. As our simulation models simulate the total number of infections, we are able to generate estimates of the undercount of COVID-19 infections in Germany (see Figure A5 in Appendix 2). According to these estimates, the real numbers during the first infection wave in spring 2020 were roughly double the size of the reported numbers, which is approximately in line with earlier research on the undercount in Germany (Bohk-Ewald et al. 2020). Thus, to gain a better understanding of how this bias might affect our outcomes, we run the model for two different scenarios: one in which we use the official incidence data, and a second scenario in which, based on our underreporting estimates, we multiply these numbers at the initial date throughout Germany by 2.

Figure 3 illustrates main outcomes of this first case. The first scenario with no adjustment to the incidence data is presented in the upper sub-figure, the second scenario with a 50% undercount assumption in the lower sub-figure. It demonstrates that with a constant R_ts_ = 3.5 there is indeed some variation across the German states when ICU capacities would have been reached. In the second scenario, the limit is reached earlier, while the spatial distribution pattern is not much different from the first scenario. As indicated before, critical situations would already be likely to emerge if the COVID-19 related demand for ICU capacities would reach a level of 35% (as up to 65% of available ICU beds are occupied by patients without COVID-19). The southern federal states Baden-Württemberg and Bavaria, which are located in close vicinity to early European epicenters such as Northern Italy and parts of Austria, would have reached critical ICU demand levels based on scenario 1 by the mid of April, while based on scenario 2 this would have happened about 7 days earlier. In both scenarios, within a corridor of about two weeks, all other states would have followed, with particularly eastern German states lagging behind (see also Figure A2 in Appendix 2). Thus, while doubling the number of infections at the initial date has some effects on the temporal pattern, it has less effect on the temporal lag between the states. In interpreting these numbers, it is relevant to note that our models assume no transfers of ICU patients between federal states. Thus, under the more realistic assumption that such relocation would happen, ICU capacities of the lagging states would have been reached a bit earlier.

**Figure 3:**
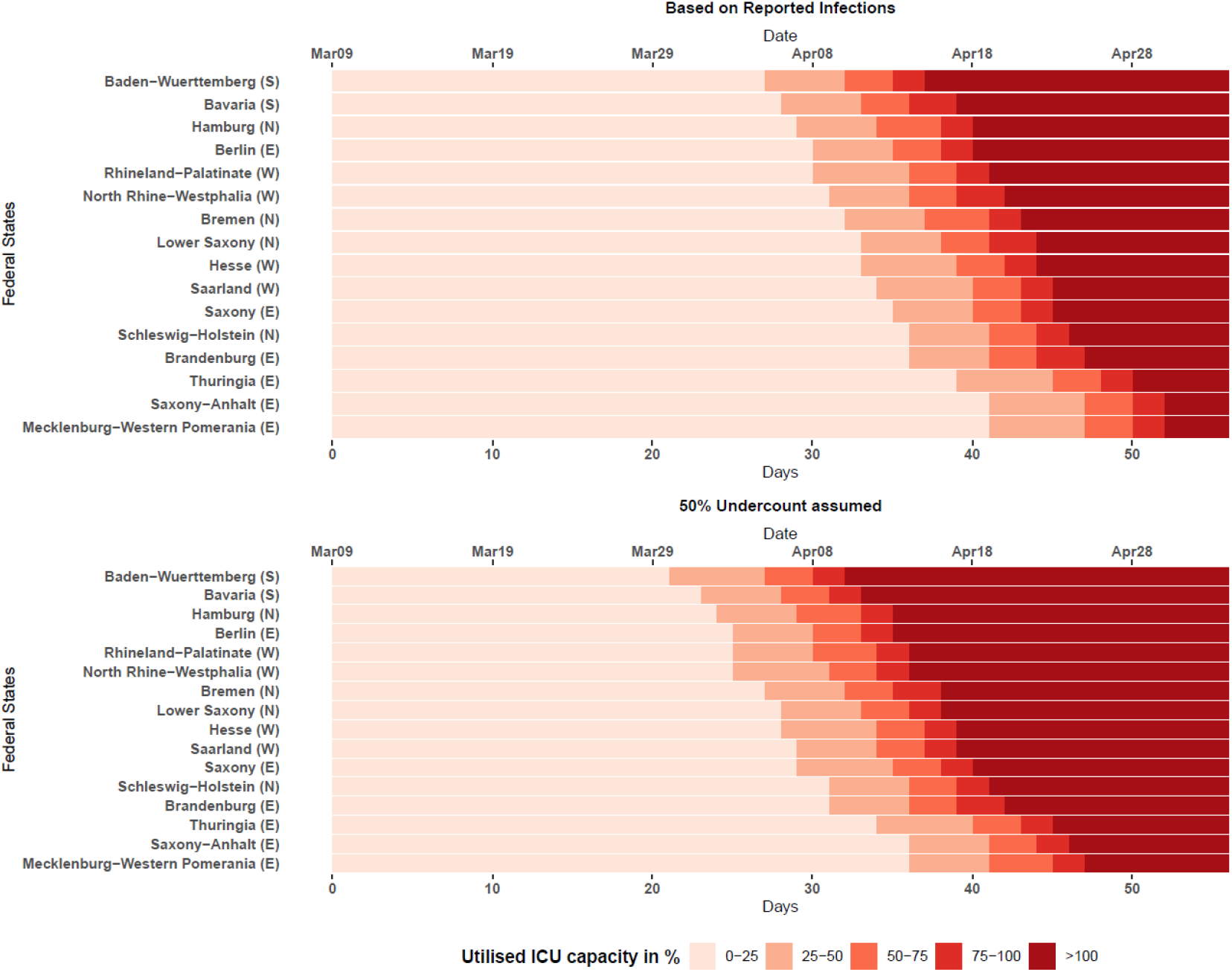
Simulated effect of an uncontained first COVID-19 wave on COVID-19-related ICU occupancy in German federal states (case 1) Note: The figure describes the simulated degree to which ICU capacities would be utilized by patients with COVID-19. Already levels above 50% should be considered critical, taking into account that 50%-65% of the ICU beds are typically occupied by patients with other illnesses. The upper figures describe scenario 1 (based on reported numbers), the lower subfigure scenario 2 (assuming an undercount of 50%). The simulation is based on the assumption that no ICU patients are transferred between federal states. If they would be transferred, capacities in lagging states would have been reached earlier. Source: RKI, German Federal Employment Agency, Federal Statistical Office of Germany, own calculations.

The take home message from this hypothetical scenario is that without any NPIs we would likely have witnessed some temporal variation in when ICU capacities would have been reached, but that the temporal windows would have been very narrow in an uncontained pandemic.

### Case 2: Assessment of NPI-level appropriateness in a low-dynamic period

In the second case, we demonstrate how our model can assist in assessing the appropriateness of existing NPI-levels for a low-dynamic period that Germany experienced during the summer months. The case presents a forecast based on data available on the 30^th^ of June. The main outcomes for all 16 federal states are provided in Figure 4 and the forecasts for the 38 NUTS-2 regions are presented in Figure A3 in Appendix 2. Before presenting the outcomes, we will use Figure 4 to briefly illustrate our forecast procedure. For this we focus in Figure 4 on the sub-figure for the city state of Berlin which consists only of one NUTS-2 region, so that the federal state level and the NUTS-2 regional level coincide in this case (for federal states with more than one NUTS-2 region the optimization is performed at a lower level of spatial aggregation). The dotted line depicts the reported ICU patients with COVID-19. Against the ICU patient trend data that was reported until the 30^th^ of June, we optimize our model to generate the *μ*_*ts*_-reductions for Berlin. The grey solid line shows the ICU trend data which we simulated with our model beginning on the 9^th^ of March up until the 30^th^ of June. It demonstrates that by adapting our *μ*_*ts*_-reductions, our model is able to replicate the ICU trend data very well.

**Figure 4:**
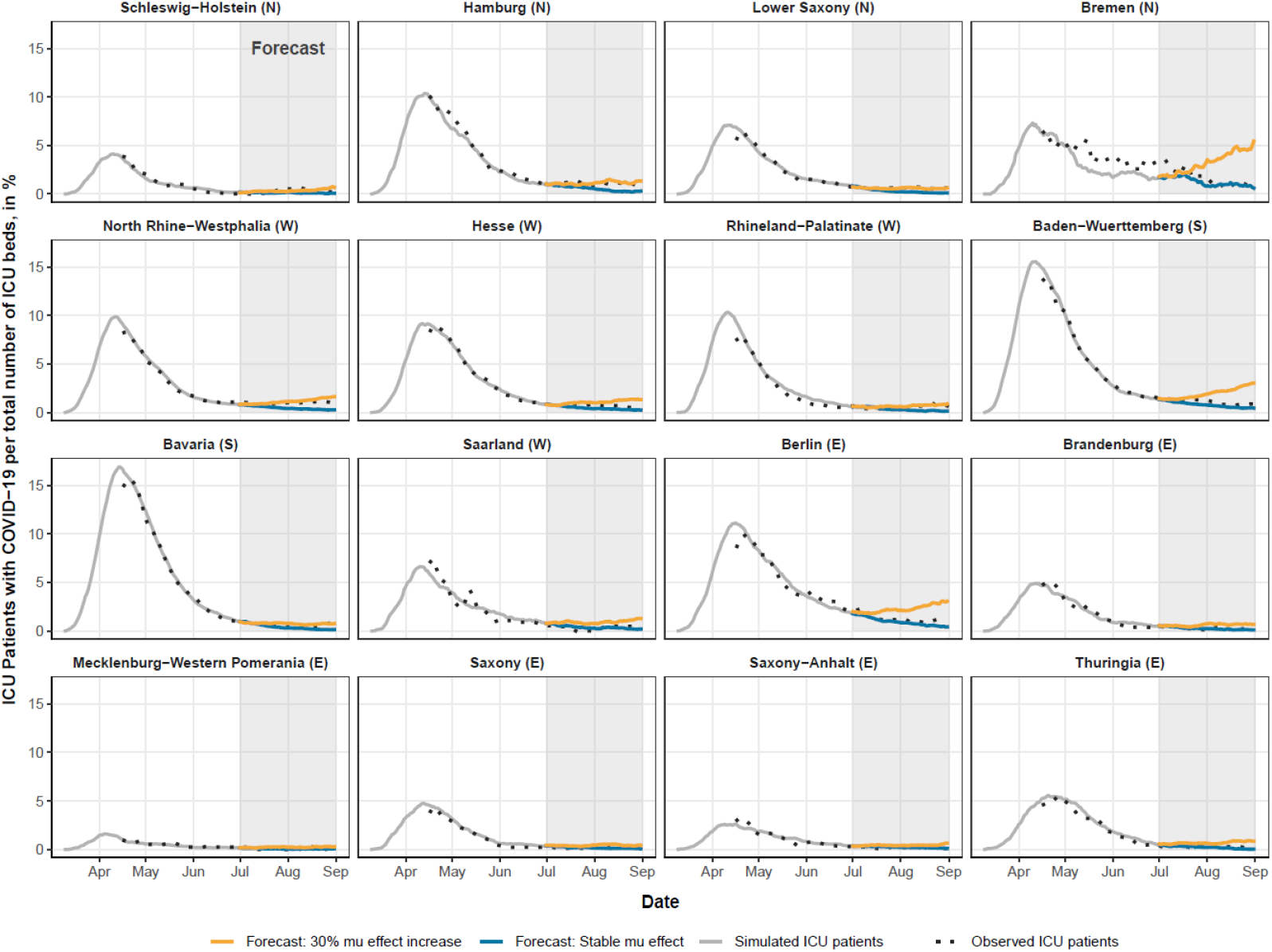
Forecast of COVID-19-related ICU demand in low-dynamic setting (case 2) Note: The figure describes the simulated degree to which ICU capacities would be utilized by patients with COVID-19. Already levels above 50% should be considered critical, taking into account that 50%-65% of the ICU beds are typically occupied by patients with other illnesses. Source: RKI, German Federal Employment Agency, Federal Statistical Office of Germany, own calculations.

We then provide forecasts for two different *μ*_*ts*_-scenarios. For the interpretation of these scenarios, it is relevant to stress one more time that these *μ*_*ts*_-scenarios are not only driven by changes in NPIs, but also by other aspects (changes in the risk profile of newly infected persons, improvements in medical treatments, seasonal effects). In one scenario (blue), we keep the latest estimated *μ*_*ts*_-effects for the NUTS-2 regions of each federal state constant for the entire forecasting period. In a second scenario (yellow), we increase the estimated NUTS-2 level *μ*_*ts*_-effect at the beginning of July by 30%. Instead of phasing this effect in, we introduce it as a “shock” at the beginning of the period, as this eases interpretation of the outcome.

Under the constant *μ*_*ts*_-effect scenario, we get flat lines with very low ICU occupancy rates of patients with COVID-19 for all states. For almost all states, this is in line with the ICU patient development that was reported in August and September. More importantly, for the 30% *μ*_*ts*_ increase scenario, we see indeed quite some variation across the states. In some states such as Bremen, Berlin, and Baden-Wuerttemberg, a 30% *μ*_*ts*_ increase would have resulted in an increase in the share of occupied ICU beds in the next 2 months to come, while in most of the other states the forecast of the 30% *μ*_*ts*_-increase scenario does not result in massive upward trends during the forecasting period. This indicates that the elasticity of different states to *μ*_*ts*_-increases, e.g. as a result of relaxing NPIs, varied at that time throughout Germany. Another finding is that in such a low-dynamic setting, a shock-like 30% *μ*_*ts*_ -increase would take between one and several months to take substantial effect on ICU demand.

### Case 3: Assessment of NPI-level appropriateness in a high-dynamic period

The third case deals with the highly dynamic resurgence of the COVID-19 pandemic in autumn 2020 (see Figure 5 for federal states and Figure A4 in Appendix 2 for NUTS-2 regions). We focus here analogous to the prior scenario on spatial variation in projected ICU demand across the 16 federal states. The forecast period starts on the 2^nd^ of November when a partial lockdown was introduced in Germany including the closure of restaurants, entertainment facilities, and public recreation centers.

**Figure 5:**
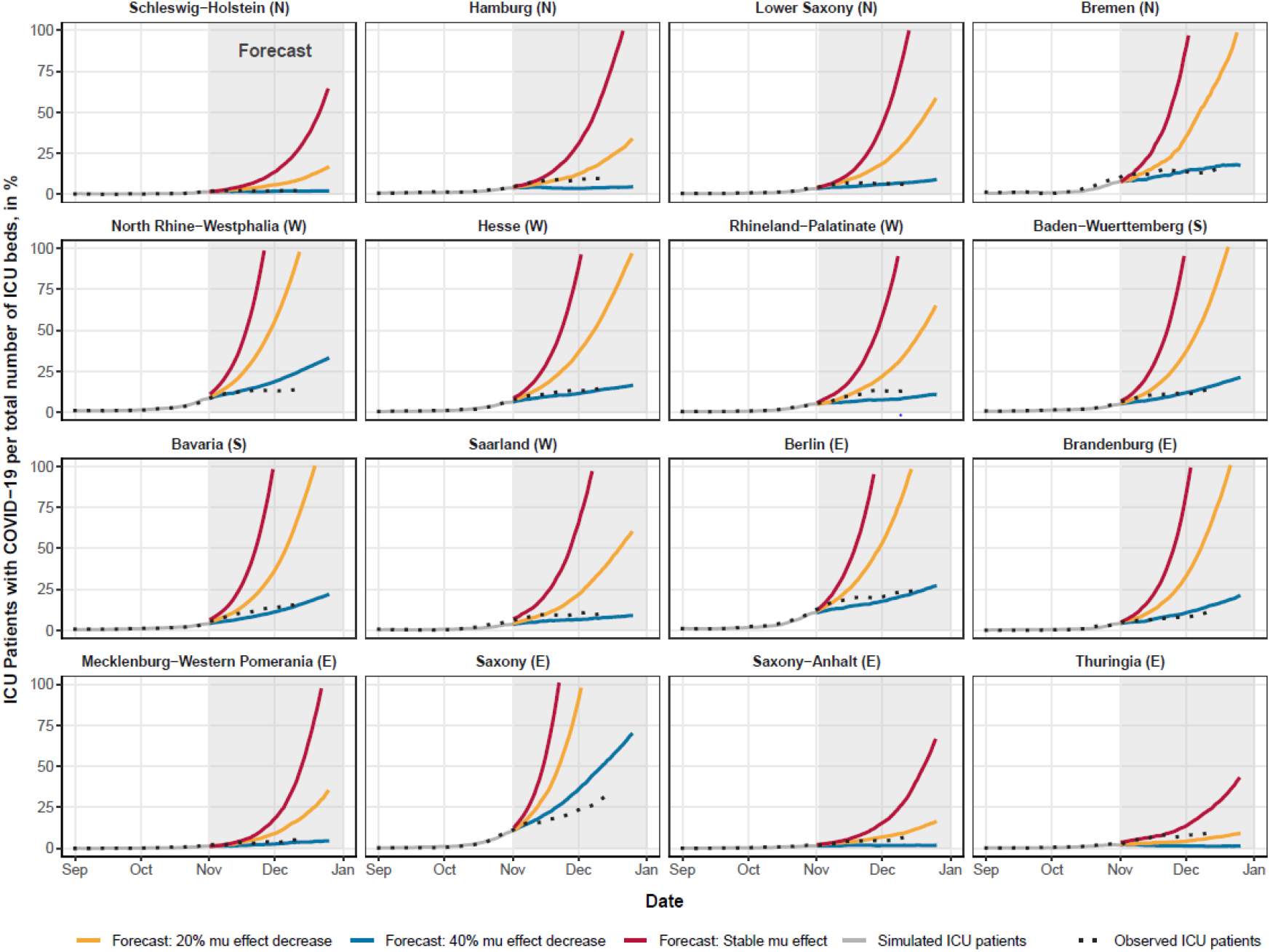
Forecast of COVID-19-related ICU demand in high-dynamic setting (case 3) Note: The figure describes the simulated degree to which ICU capacities would be utilized by patients with COVID-19. Already levels above 50% should be considered critical, taking into account that 50%-65% of the ICU beds are typically occupied by patients with other illnesses. Source: RKI, German Federal Employment Agency, Federal Statistical Office of Germany, own calculations.

In this case, we introduce three different scenarios. In the first scenario, we keep the last obtained *μ*_*ts*_-value for each federal state constant to see how the epidemic would have had evolved with no adjustments to *μ*_*ts*_. In the second and third scenario, we simulate how a lockdown-imposed immediate reduction of *μ*_*ts*_ by 20% and 40%, respectively, on the 2^nd^ of November would have affected ICU demand in the months to come.

In the first scenario with a constant *μ*_*ts*_, the forecasted COVID-19-related ICU demand in most federal states would reach more than 100% of the overall ICU capacity levels by the beginning of November. Accordingly, our model suggests that in this highly dynamic setting all federal states should aim to reduce their *μ*_*ts*_-values in order to keep the pandemic manageable. Following the forecasts of the second and third scenario, there is hardly any variation in the degree to which different states would need to implement additional NPIs. Only in very few states in Northern and Eastern Germany, *μ*_*ts*_-reductions by 20% would flatten the curve and would keep the COVID-19 related ICU demand below 25% of the overall ICU-capacity levels. A *μ*_*ts*_ -reduction by 40%, however, succeeds in keeping the number of ICU patients with COVID-19 below 25% of the overall ICU-capacity levels in most federal states. Nevertheless, even a 40%-reduction does not result in decreasing numbers of ICU-patients by the end of the year, which is particularly true for the heavily affected state of Saxony. The simulations additionally show that the 40%-decrease in *μ*_*ts*_ follows the observed number of ICU-patients reasonably well, indicating that the partial lockdown imposed on the 2^nd^ of November 2020 reduced social contacts by up to 40%, ceteris paribus.

To summarize, according to our simulations, in this highly dynamic setting, substantial additional *μ*_*ts*_ reduction seemed to be necessary in all federal states to avoid the overburdening of the ICU health care system. In contrast to the low-dynamic setting, there was hardly any room for variation in NPIs across federal states in the high-dynamic period. Finally, our forecasts predict that it would take at least 2 to 3 weeks in high-dynamic settings until the introduction of partial lockdown measures would decelerate the increase in the number of ICU patients.

## Discussion and Conclusion

This paper presented a microsimulation model of the COVID-19 pandemic which aims to assist policy and administrative decision makers in assessing whether the current level of NPIs should be maintained or even strengthened to prevent ICU demand from exceeding supply, or whether there is leeway to relax them. We have demonstrated how our model can be used for such assessments in different settings. The outcomes for a low dynamic situation identified some variation in potential for relaxing NPIs across the federal states. In the observed period with high dynamic, on the other hand, our model indicates that all federal states should strengthen their NPIs in order to keep the pandemic at a manageable level. In addition, in the high-dynamic period we observed hardly any variation on the federal state level in the degree to which additional NPIs seem to be needed. Moreover, the models’ outcomes suggest that in large infection waves as experienced in Germany in spring and autumn, ICU demand would soon exceed supply if no containment measures were enacted.

Limitations of our approach include that our reduction factor *μ*_*ts*_ captures any kind of factor that affects the infection risks, including also effects that cannot be controlled by policy makers or societies such as seasonal changes in weather conditions. However, further refinements of the models would offer the possibility to isolate some of these factors. Over the course of the pandemic we might obtain a better understanding of the seasonal effects, and could incorporate them into the forecasts. Reduced COVID-19 related ICU demand due to improvements in medical treatments could be accounted for by adjusting our epidemiological parameters over time, based on data from epidemiological studies. If the quality of incidence data further improves as the result of increased test capacities, we might also be able to better account for how infection intensities vary by age across sub-national regions at a given time.

While focusing on ICU coverage offers the advantage that we are able to work with informative data, one potential drawback is that shifts in the COVID-19 dynamic become visible only somewhat later in the ICU occupancy than in the incidence data. This is not necessarily a disadvantage. Over the summer, Germany had experienced a number of surges in slaughterhouses among rather young healthy employees, which created local hot spots in the incidence data, but had little effect on the ICU data. Thus, focusing on the incidence data would have likely created too high estimates of ICU demand. However, in periods with substantial change, such as in autumn 2020, our forecast can only capture the trend change in the level of infections if it is already visible in the ICU data.

Another limitation of using ICU capacities is that these are not only linked to available beds. Even if enough beds were available, there might be shortages in ICU personnel. However, our simulations demonstrate that it is a risky undertaking anyway to allow the pandemic to reach such a dynamic that high shares of the ICU capacities would be occupied by patients with COVID-19, as in such dynamic situations COVID-19 related ICU demand could quickly move to even higher levels (see cases 1 and 3). This speaks in favor of aiming to preemptively contain the pandemic at lower levels to ensure that the pandemic is not at risk of running out of control, and that also the substantial demand for ICU beds independent of COVID-19 can be met. Another limitation is that *μ*_*ts*_-effects are also affected by the degree to which the local health affairs offices can isolate infected persons and trace their contacts. This is likely better possible at low COVID-19 dynamics than at high COVID-19 dynamics, while our model implicitly assumes this effect to be constant and independent of the dynamic.

In addition, it is also debatable whether the commuter matrix is best suited to identify spatial pathways of the diffusion. Commuter matrices do, for example, not account for touristic movements. In this regard, movement data from cell phones might have more appeal (Schlosser et al. 2020). However, our model would allow the incorporation of such data. But as we did not have such data available, we decided to use a two-fold strategy to account for this. One is the use of a commuter matrix as a proxy for connectivity. In addition, we apply a parameter *(w_obs*) to redistribute infection cases simulated by our model based on the regional distribution of the incidence data for the period prior to the forecast (see Appendix 1). This also allows us to account to some degree for spatial diffusion pathways. A final limitation is that the catchment areas of hospitals vary. University hospitals especially get patients from further away. This is of relevance for our city states estimates and when accounting for variation at lower geographical levels of aggregation, as they affect the distribution of the infected cases that we generate. Also in that regard, the *w_obs* parameter allows us to counteract this biasing effect to some extent.

The models presented in this paper attempt to account for uncertainty about the future disease propagation through different *μ*_*ts*_-scenarios. In a future extension, we aim for alternative strategies to take statistical uncertainty into account. However, related to this we face the limitation that in a dynamic pandemic uncertainty can derive from many different sources (policy changes, data limitations, model assumptions, dynamics in neighboring countries, seasonal effects, virus mutations, new vaccinations etc.). Next to the cases presented in this paper, the model could also be used for other purposes. This includes, for example, hypothetical scenarios with regional and temporal variation in lockdown measures. Our model would allow users to implement such variation at the district level.

The model is implemented in R, and all code and the data used for the simulations are freely available online in order to make our results replicable. The code can be applied to countries other than Germany, provided that reliable data on the number of ICU patients infected with COVID-19 and data on spatial proximity are available at sub-national level. In parallel, we are also working on a Fortran version of the model, which would allow the simulation to work with 100% samples of populations.

## Data Availability

The data that support the findings of this study are openly available.

## Acknowledgements

The authors would like to thank Jakub Bijak, Andreas Ruopp, Martin Schumacher, Frans Willekens, and Sabine Zinn for comments on earlier versions of the paper. We also express gratitude to Harun Sulak for support on one of the figures.

## Appendix 1: Data and Methods

### Data

In monitoring the spatial dynamic of the pandemic in Germany, we use as input for our microsimulations two data sources. From the first data source we derive daily updated data on (1) the number of intensive care unit (ICU) patients diagnosed with COVID-19, (2) available ICU beds, and (3) deaths of persons with COVID-19 in an ICU by day and district. These data are provided by the German intensive care register maintained by the German Interdisciplinary Association for Intensive Care and Emergency Medicine (DIVI) and the Robert Koch-Institute (RKI). We use both an open access dataset (RKI 2020a) as well as a more detailed restricted access dataset to which we have access through our RKI collaborators (RKI 2020b). The second data source maintained by the Robert Koch-Institute gives access to daily updated statistics on the registered COVID-19-case incidences and deaths of persons with a COVID-19 infection by district, sex and broad age groups (0-4, 5-14, 15-34, 35-59, 60-79, 80+) (RKI 2020d).

The German intensive care register is a new data source which was established in spring 2020 as part of a strategy to improve access to data with high relevance for monitoring the COVID-19 pandemic. Since the middle of March, a growing number of German hospitals with ICUs voluntarily participated in reporting the number of patients diagnosed with COVID-19 requiring intensive care on a daily basis. Reliable data on ICU occupancy of persons with COVID-19 covering the entire country are available from mid-April onwards when participation in the registry became mandatory for all German hospitals with ICUs. The intensive bed register is maintained by the intensive care departments of the hospitals. The IT infrastructure of this register is designed in such a way that daily statistics on ICU patients with COVID-19 can be collected with high reliability, which is one of the main motivations why we calibrate our microsimulation against these data. Data on the number of deceased persons with COVID-19 in an ICU, which is available in the dataset with restricted access (RKI 2020b), are used for assessments of the share of COVID-19 deaths occurring in an ICU.

For our model, we mostly rely on publicly available data by district, which were first published on April 30, 2020. We complement these data with a data series from April 15 to April 29, 2020 from the restricted-access dataset (RKI 2020b). These data on ICU beds by day and hospital were also used to implement some data corrections (see below). That reliable ICU data are only available from 15^th^ of April 2020 onwards provides us with a challenge when capturing the dynamic of the first COVID-19 wave Germany experienced in spring 2020, as in some states the number of ICU patients with COVID-19 was already falling in mid-April.

We treated the received ICU occupancy data of persons diagnosed with COVID-19 as follows: For the period up until April 29, when a certain hospital made several entries on a specific day, the entry with the highest number of ICU patients with COVID-19 was chosen. When a certain hospital did not make any entry on a specific day, the mean number of ICU patients entered on the day before and the day after was chosen. From April 30 on, we rely on the district-level statistics as available in the open-access data (RKI 2020a). To this dataset, we only applied one correction as identified in the restricted-access dataset (RKI 2020b): One hospital in Saxony provided implausibly high COVID-19 statistics prior to August 5, 2020 when the number of ICU patients suddenly dropped from a total of 23 to a total of 4. For that reason, we decided to exclude this specific hospital prior to that date, as we would otherwise have an artificial downward jump in the ICU data series of Saxony. No corrections were applied to the total number of available ICU beds and the ICU deaths of patients with COVID-19.

As we explained in the introduction of our paper, our forecasts are mostly based on the very reliable ICU data. Nevertheless, for some purposes we also fall back on incidence data and on statistics on the number of deaths of persons with a COVID-19 infection as provided by the RKI (2020d). At the beginning of the COVID-19 pandemic, Germany already had a quite well-developed system of testing capacities in place throughout the country. This enabled the country especially at the beginning of the pandemic to capture COVID-19 infections to a higher degree than many other countries (Stang et al. 2020b). Nevertheless, also in Germany many cases remain unreported for different reasons (Streeck et al. 2020; Bohk-Ewald et al. 2020). Related to our aim to adequately capture the temporal dynamic of the pandemic, it is important to note that for the incidence statistics the RKI aims to collect not only the date at which a case was registered with the local health authorities (*ger: Meldedatum*), but also the so-called reference date (*ger: Referenzdatum*) at which according to self-reports of the infected persons the first symptoms occurred. While all case entries have a date of registration, the date when first symptoms occurred is not available for all registered cases. For those cases for which the day at which first symptoms occurred is unknown, the RKI uses as reference date the date of registration. There can also be cases in which the date of registration is actually prior to the date at which first symptoms occurred. These are not necessarily data errors, as sometimes contact persons of persons with a positive test are entered in the system independent of whether they have already symptoms and/or a positive test (an der Heiden & Hamouda 2020). If these people develop a reported COVID-19 infection after the first registration, the date of registration is prior to the date at which first symptoms occurred. It is also relevant to note that the RKI and local health authorities are striving to fix data issues for older cases. Thus, it can happen that cases which first only provide the date of registration receive the date when first symptoms occurred added with a delay of days or several weeks.

For our attempt to capture the temporal dynamic, the reference date is more accurate. We therefore decided to impute reference dates for those cases for which only the registration date is available. This imputation we apply on a daily base, using the latest available data. We impute the beginning of the illness for the cases for which we only know the date of registration, using by day the distribution of the delay in the correct cases between the date of reference and the date of registration (for whole Germany) as a reference. We illustrate this with a simple example, in which on one day ten cases were registered in Germany, of which four lack the reference date. Of the remaining six cases, three were registered with a three-day delay since the first symptoms occurred, two with a two-day delay, and one with a one-day delay. From this reference distribution, we derive the cumulative proportions (0.5, 0.83, 1). We then sample from a uniform distribution between 0 and 1 four numbers to impute the delay for the cases without a reference date (e.g., if we draw a 0.4, the imputed delay would be three days). The RKI seems to use a similar approach, though they do it separately by age and sex (an der Heiden & Hamouda 2020), but not by day. We decided to impute by day, as the difference between the date of reference and date of registration is likely to be larger if there is a weekend in-between, when many local health authorities are not registering cases, than in cases where the date of reference and the date of registration were reported in the same week. Therefore, our correction approach seems better suited to account for regular weekly variation in reporting.

The cumulative data on COVID-19 deaths by different levels of spatial aggregation also has some shortcomings, so that we only use it for consistency checks. For the deaths of persons with a COVID-19 infection, we face the problem that for data confidentiality reasons the RKI does not provide the exact date of death. Instead, it provides for these cases the date when the COVID-19 infection of the deceased person was registered (*ger: Meldedatum*) for the first time and when this person reported first symptoms (if available, ger: *Referenzdatum*). To apply corrections, we contrast the data by registration date with correct aggregate-level data on deaths of persons with COVID-19 by day, which for Germany is unfortunately only available at the national level, and not for the spatial detail which we would require. This provides us with an indication that there are on average 6 days between the death date of a person and the date at which this person was first registered as having COVID-19. We thus add 6 days to the reference date in an attempt to approximate the date of death. This rather crude adjustment is justified in our eyes, as we (1) focus on cumulative death trends, which are less affected by small temporal inconsistencies, and as (2) we use these data only for cross-validation.

As the spread of the virus in Germany differs spatially, we introduce population data on the district level, considering each district’s current population-age distribution. The age structure varies considerably between German districts and plays an important role in the current pandemic because elderly people and those with existing co-morbidities are much more likely to be severely affected by the disease (Dowd et al. 2020; Dudel et al. 2020; Ioannidis et al. 2020; Nepomuceno et al. 2020). Data on the total population count per district broken down by 5-year age groups up to age 90+ is derived from the Federal Statistical Office of Germany (*Statistisches Bundesamt*). We use the most recent available population data for December 31, 2018 (we intend to switch to data for the end of 2019 as soon as it becomes available).

An additional feature of our simulation model is that the risk of susceptible individuals to contract COVID-19 at time point t_n_ depends on the regional distribution of the contagious population at time point t_n-1_ as well as on the general connectivity between the 401 German districts. As an indicator for the district connectivity, we use data on commuting behavior provided by the German Federal Employment Agency (*Bundesagentur für Arbeit*). These data contain aggregated information on the district of residence and on the district of workplace for all employed individuals who are subject to social insurance contribution (more than 33 million employees in total, who represent about three-fourths of the working population in Germany). Based on these commuting data we are able to construct the connectivity between all 401 German districts in a connectivity matrix. First, we constructed a symmetric matrix with the total number of persons commuting between two districts in each off-diagonal matrix element, and the total number of employees whose workplace and place of residence are located within the same district on the matrix diagonal. Second, we derive row-wise proportions. Accordingly, each matrix row adds up to 1 (100%).

An assessment of changes in social contact frequency, an important aspect of NPIs, is derived from an online survey of the Max Planck Institute for Demographic Research (Perrotta et al. 2020; Del Fava et al. 2020). It provides weekly information in social contact frequency by country, age group and type of contact (e.g. in households and outside of the households). These changes in social contact frequency provided by the MPIDR were used to determine initial parameter search spaces for calibrating *µ*_*ts*_, and for cross-checks of the obtained *µ*_*ts*_ values.

### Methods

With exception of example case 1, which was calculated with 100% of the German population, simulations are run for a 20%-sample of the current German population (16,603,842) on the high performance computing system Hawk which is equipped with AMD EPYC™ 7742 processors. Depending on the simulated timeframe each model execution with a 20% sample of the German population requires 2 GB - 6 GB of main memory. The microsimulations have been programmed in R 4.0.2. In addition to functions available in the pre-installed packages, efficient data structures and functions from the packages *dplyr, rlist*, and *data*.*table* were applied. The calibration procedure is implemented based on the genetic algorithms provided by the *GA* package, whereas for the parameter space exploration functions provided by the *lhs* package were applied. For parallel deployment, the *doMPI, parallelly* and *doParallel* packages were used along with the *doRNG* package for parallel seeding. The package *ggplot2* along with functions from the packages *grid, gridExtra, pracma* and *RColorBrewer* are used to visualize the simulation results. All case studies presented in this paper rely on microsimulations starting on March 9, 2020 (see main paper for the motivation).

Figure A1 shows the three levels of spatial aggregation used in our simulation model. We run our simulation model at the NUTS-3 level of the 401 German districts and our simulation sample is assigned to the districts based on data on the districts’ population size and age structure as derived from the Federal Statistical Office of Germany (2020). The transition from (1) susceptible to (2) latently infected occurs at the district level as well. Whether an individual residing in district *i* gets infected depends on the share of infectious persons in the residential district *i* and, if the parameter *w_int* is unequal 1, the share of infectious persons in neighboring districts *j* with which district *i* is socially connected. As proxy for social connectivity we use commuter flow data derived from the German Federal Employment Agency (2019).

The calibration against ICU data is applied on the level of the 38 NUTS-2 regions of Germany. We opted for a calibration on a higher level of spatial aggregation as many districts have small or no ICU capacities at all, so that severely infected patients frequently have to be diverted to neighboring districts with for instance university medical centers. Finally, we aggregate the simulation outcomes to the level of the 16 federal states (NUTS-1) to increase the clarity and readability of the results.

**Figure A1:**
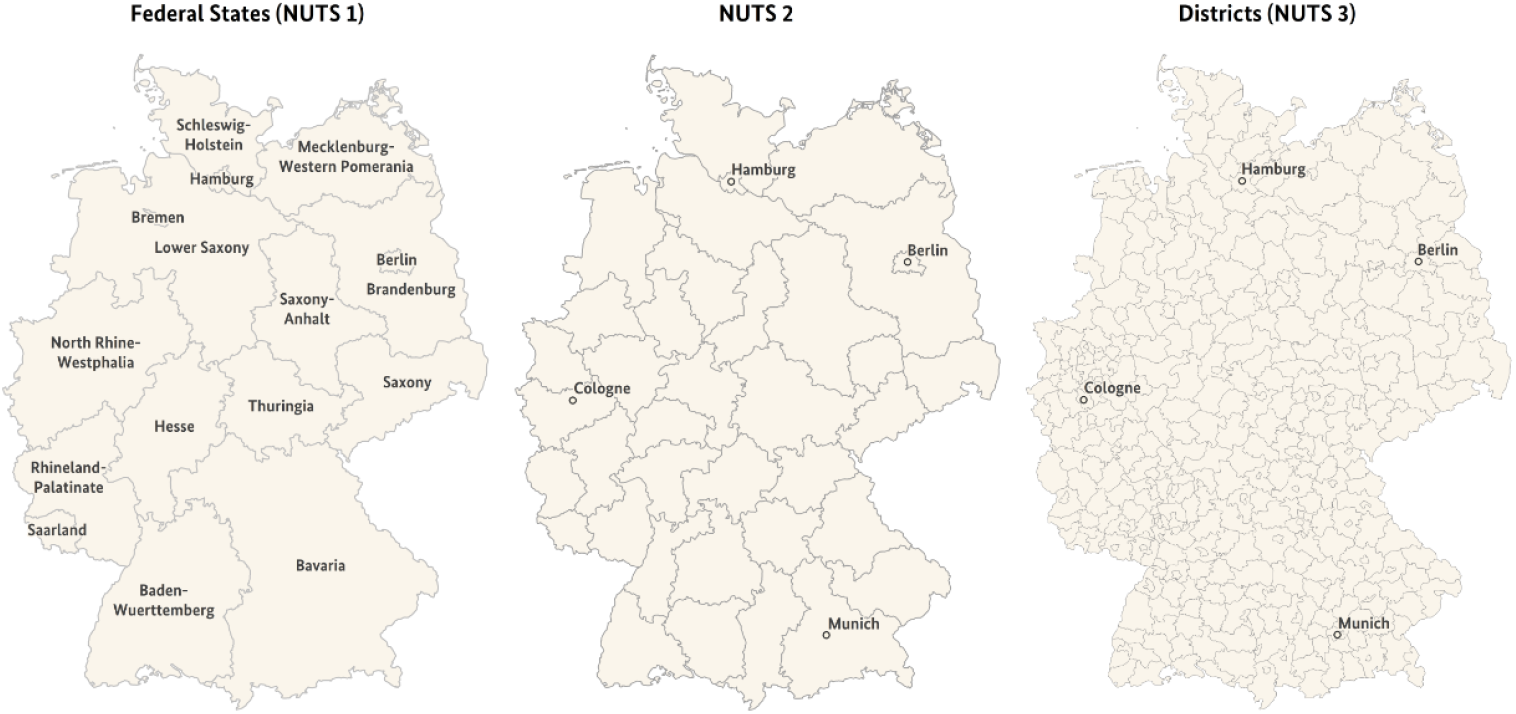
The three levels of spatial aggregation used in our simulation model. *States, transitions, and parameter specifications*

An overview over all parameters is provided in Table A1.

Following an extended SEIR-model, individuals in our microsimulation can obtain seven distinct states: (1) Susceptible; (2) Latently Infected; (3) Infected with COVID-19 and contagious; (4) Illness and contagious; (5) Death; (6) Intensive care unit (ICU); and (7) Recovered (see Figure 1). The infection risk (transition from state 1 to state 2) of an individual at time *t* in federal state *s* is determined by two aspects: (1) the time-varying reproduction number *R*_*ts*_ and (2) the share of contagious people in the spatial surrounding. How we derive the reproduction number *R*_*ts*_ through calibration is detailed in the methodological section of our paper, while additional details on the calibration process are provided below in the section calibration.

We attempt to approximate the share of contagious people in the spatial surrounding in the base specification of the model with the share of contagious persons in the residential district. As an additional option, the model controls for spatial diffusion pathways due to mobility of humans between districts with the *w_int* parameter. When this information is taken into account, the infection risk of an individual living in district *d*_*i*_ is not only affected by the level of infections in district *d*_*i*_, but to some degree also by the level of infections in neighboring districts *d*_*j*_ with which district *d*_*i*_ has commuter flow relations. The parameter *w_int* can vary between 0 and 1, with e.g. 0.9 implying that the infection risk is to 90% dependent on the share of infected persons in district *d*_*i*_, and to 10% dependent on the commuter-flow weighted share of infected persons in districts *d*_*j*_. Sensitivity checks suggest that a value or 0.9 seems to approximate diffusion effects quite well.

The transition from state 1 (susceptible) to state 2 (latently infected) is illustrated in equation (1) where *S*(*t, i*) is the susceptible population in district i at time t, *I*(*t, i*) is the infected population in the contagious state 3 in district i at time t, *J*(*t, i*) is the ill and contagious population (state 4) in district i at time t, and N(i) is the total population in district i. *b*_*I*_ and *b*_*J*_ are the infection rates for state 3 and state 4 respectively, which might be identical. *P*(*i, j*) represents the specific entry of the commuter matrix P for districts i and j, and *w_int* is the parameter described above.

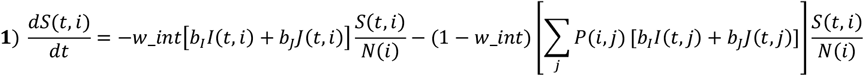

If in the simulation a person becomes infected (i.e., moves from state 1 to state 2), the subsequent transitions to state 3 and state 4 are occurring in a deterministic manner that is identical for all individuals. This simplification is motivated by the fact that we have no indications that these periods differ in a systematic manner across space and time. Model parameters allow variation in the length in days for which individuals stay in states 2 and 3. In line with estimates provided by an der Heiden and Buchholz (2020), pre-symptomatic individuals remain in state 2 (infected without symptoms, noncontagious) for three days after virus infection and are in state 3 (infected without symptoms, contagious) on the fourth and fifth day. Five days after the initial infection, individuals enter the state 4 illness (symptomatic or asymptomatic, contagious). These transitions are illustrated in the following equations where *E*_*k*_(*t, i*) is the exposed population (state 2) on the k-th day of the exposure and n is the last day in state 2 (e.g., 3rd day), or last day in state 3.

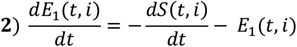

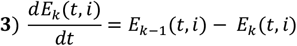

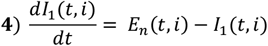

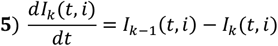

Beginning with the fifth day of infection, persons are considered ill and are at risk of dying due to a COVID-19-infection. The transition to death depends on the individual’s age (see table A1 for the age-specific transition risks). The population mortality risk is estimated at 1.6% of all infected individuals. After eight days of illness, individuals either enter the healthy/immune state or are admitted to ICU (an der Heiden & Buchholz 2020). This in turn is a simplification, but tests with more complex specifications in which we varied the transition risks over the days of the illness did not yield substantially better model results. Following Stang et al. (2020a), we assume that around 3% of the infected population is admitted to ICU. As in the case of mortality, this transition probability depends on the individual’s age (see table A1 for the age-specific transition risks). ICU patients are continuously at risk of dying and based on the ICU data provided by RKI we assume that 30% of those admitted to ICU eventually die (RKI 2020b; see also Nachtigall et al. 2020; Karagiannidis et al. 2020). We do not assume any age gradient in the transition from ICU to death. After 14 days, ICU survivors are released from ICU and considered healthy/immune. Currently, we assume that healthy individuals remain immune throughout the entire period, but the model allows to lift this restriction. If this option is chosen, individuals return at the end of their illness to state 1 (healthy, not immune). These final transitions are illustrated in the following equations where *C*(*t, i*) is the ICU population in district i a time t, *D*(*t, i*) is the deceased population in district i at time t, and *R*(*t, i*) is the recovered population in district *i* at time *t. c* represents the risk of moving to ICU if ill, *d*_*J*_ the risk of dying if ill, *r*_*J*_ the risk of recovering if ill, *r*_*C*_ the risk of recovery if in ICU, and *d*_*C*_ the risk of dying if in ICU.

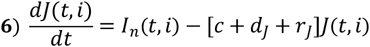

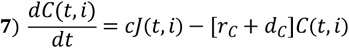

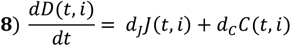

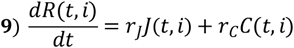

As we apply the optimization presented below at the level of the 38 NUTS-2 regions, this limits our ability to account for newly emerging hot spots at lower levels of spatial aggregation. In order to keep our model “on track” regarding the spatial pattern at the district level, we implemented the optional parameter *w_obs* that allows us for the optimization period to redistribute the simulated infections at time *t* in the districts of NUTS-2 region *s* based on district-level proportions of reported incidence data for that time *t*. The parameter can vary between 0 and 1. In the cases presented here, *w_obs* was set to 0.5, which implies that when determining the spatial distribution of new infections across the districts at day *t*, we take to 50% the spatial distribution of infections generated by the model into account, and to 50% the registered distribution based on incidence data. This redistribution assumes that the incidence data provides reliable enough information on the spatial distribution of infections, an assumption that for Germany with its well-developed testing facilities is in our eyes justified. In other countries, *w_obs* should probably be used with more caution. Sensitivity checks show that for the state level outcomes presented in this paper, the effect of *w_obs* is rather limited. Finally, we assume that individuals who developed symptoms are assumed to be 30% less contagious than pre-symptomatic individuals. This assumption allows us to control for the fact that ill persons with symptoms are more likely to get tested, to become aware of their disease and to self-isolate than those who have not developed symptoms yet. Moreover, it incorporates research on COVID-19 indicating that a substantial share of transmissions is occurring at a pre-symptomatic stage (He et al. 2020; Ganyani et al. 2020).

### Calibration process

Due to the large number of parameters in the model which are subject to calibration, for the initial calibration process, we applied a stepwise approach based on a compromise between automatic optimization and manual intervention. For the initial 570 model parameters *µ*_*ts*_ with *t ∈ [1,15]* and *s ∈ [1,38]* (for each NUTS-2 region), which covered the first wave of ICU cases until end of June 2020, i.e. the first 15 weeks in the simulation timeframe, this methodology proved to be the most time efficient approach. One limitation of the calibration in the initial period of the epidemic is that ICU-data on patients with COVID 19 is only available from the 15^th^ of April on, while we start our model on the 9^th^ of March. This implies that we cannot exactly calibrate our model for the period between the 9^th^ of March and the 15^th^ of April – the calibration just attempts to optimize each *µ*_*ts*_ level in such a way that the simulated values match the observed values from mid-April on. Thus, the simulated trends in this earliest period prior to mid-April should be rather considered as a “burn-in phase”.

The applied methodology consisted out of two steps. In each of these steps an automatic optimization by means of the *GA* package in R (Scrucca 2013) was applied in order to determine *µ*_*ts*_, so that the model results would reproduce the observed ICU cases on a certain level of aggregation. In the first step, a set of *µ*_*ts*_ parameters for the 16 federal states forming the NUTS-1 regions of Germany was determined. In the second step each NUTS-2 region was assigned the *µ*_*ts*_ parameter set of the NUTS-1 region of which it is part of (the NUTS categorization is hierarchical). Based on the NUTS-1 solution as an initial guess, a set of *µ*_*ts*_ parameters for the NUTS-2 level was determined again by automatic optimization. In both steps, a piecewise calibration of parameter sets of at-most 15 weeks at NUTS-1 level and eight weeks at NUTS-2 level was implemented in order to keep the number of parameters on a manageable scale and speed up the convergence of the optimization procedure. Experience showed that it is necessary to account for the delayed appearance of ICU cases compared to the appearance of infections driven by *µ*_*ts*_, by overlapping the timeframe subject to optimization by at least 2 weeks. This means a reasonable sequence of intervals for *t* for three executions of the procedure was found to be *t*_*1*_*∈[1,8], t*_*2*_*∈[7,14], t*_*3*_*∈[13,20]*. To further speed up the search for an optimal solution, narrowed *µ*_*ts*_ parameter bounds were determined by evaluating parameter sweeps in which all *µ*_*ts*_ parameters for the next interval were synchronously altered approximately ±40% in steps of 10%.

The GA package performs stochastic optimization of an objective function by means of genetic algorithms. This means it builds a set of candidate solutions called a population whose individuals are iteratively evolved based on the principles of heredity. In order to distinguish between the model population and the population of the optimization procedure we will call the optimization procedure’s population GA-population in the following. Reasonable convergence could be achieved with 256 individuals, i.e. candidate solutions in the GA-population. The evaluation of each candidate solution, consisting out of 4-10 model evaluations covering 10 weeks took approximately 30 min. Due to hierarchical parallelization of the procedure by means of the internal parallelism of the *GA* package in combination with the *parallelly* package and the implementation of a checkpoint restart procedure avoiding unnecessary evaluations of the already calibrated weeks, the execution time for one evaluation of a GA-population could be kept almost constant, independently from the number of its candidate solutions and the progress along the sequence of *µ*_*ts*_ intervals.

Since experience showed that after 50 generations i.e. iterations no more fast convergence could be achieved, the solution was assessed and restarted with narrowed parameter bounds if needed. As already stated in the main section of this paper the root mean square of the daily distances between the observed data and the mean result of multiple model executions was used as the objective function. In order to assess the quality of the found solution on a more fine-grained level, the root mean squares of the daily distances between the observed data and the mean of the simulation results per week are used. Additionally, the root mean squares of distance between the minimum and maximum result per week are evaluated. In general, the target is to keep the root mean square per week below 10% of the mean ICU-cases per week. If this cannot be achieved, which we sometimes experienced in high dynamic situations, the second criterion is to keep the root mean square of the differences between the observed data and the average result below the root mean square of the difference between the minimum and maximum result.

We believe that based on these two criteria the described procedure can be automated in order to enable daily forecast updates, although it is not yet implemented.

**Table A1:** Parameter specification and sources.

| Parameters |  |  |  |
| --- | --- | --- | --- |
| Parameter | Object name in code | Value | Source(s) |
| Basic reproduction number $R_0$ | $R_0$ | 3.5 | **RKI 2020c** |
| Extent to which persons in state “ill, contagious” are less contagious compared to persons in the state “infected, contagious” (1.0 is reference level) | *less_contagious* | 0.7 | This parameter allows persons in the state “ill, contagious” to be made less contagious as these people are more likely to self-quarantine. In the models presented in this paper, we set this parameter to 0.7. |
| Sample size | *sam_size* | 16,603,842 (20% sample of the German population) | Federal Statistical Office of Germany (Population by age, sex and district, 31.12.2018) |
| Sample proportion (based on officially registered COVID-19 cases in Germany (1.0 is reference level)) | *sam_prop* | 1.0 (in scenario 2 of case study 1: 2.0) | Allows adjustment of incidence numbers (**RKI 2020d**) at the starting date of the model based on assumptions on the undercount. |
| Weighting parameter that assumes values between 0 and 1; with 1 implying that new infections generated by the model are redistributed across regions to match the observed distribution of registered new infections, and 0 implying that the distribution of infections across regions is determined only by the model. | *w_obs* | 0.5 | Specification derived on the basis of parameter space exploration |
| Weighting parameter that assumes values between 0 and 1; with 1 implying only within-region spread, and 0 allowing for inter-region spread based on commuter flows as described in the methods section | *w_int* | 0.9 | Specification derived based on parameter space exploration |

Table A1: Parameter specification and sources
| Duration parameters |  |  |  |
| --- | --- | --- | --- |
| Parameter | Object name in code | Value | Source(s) |
| Duration from infection to being contagious (period of latency) | *inf_dur* | 3 days | an der Heiden & Buchholz 2020 |
| Duration from being contagious to being ill (i.e. being symptomatic) | *cont_dur* | 2 days | an der Heiden & Buchholz 2020 |
| Duration from illness to immunity or entry into ICU | *ill_dur* | 8 days | an der Heiden & Buchholz 2020 |
| Duration from being at ICU to immunity | *icu_dur* | 14 days | Stang et al. 2020a<br>Karagiannidis et al. 2020 |
| Transition probabilities |  |  |  |
| Parameter | Object name in code | Value | Source(s) |
| Risk of being sent to ICU once ill, by age-group (ill, contagious state to state ICU) | *icu_risk* | 00-55: 0.000 | Stang et al. 2020a;<br>own calculations based on Germany's age structure |
|  |  | 55-60: 0.004 |  |
|  |  | 60-65: 0.008 |  |
|  |  | 65-70: 0.032 |  |
|  |  | 70-75: 0.064 |  |
|  |  | 75-80: 0.120 |  |
|  |  | 80-85: 0.200 |  |
|  |  | 85-90: 0.320 |  |
|  |  | 90+: 0.561 |  |
|  |  | **total: 0.030** |  |
| Survival chance once ill, by age-group (ill, contagious state to state healthy) | *surv_ill* | 00-55: 1.000 | Own calculations based on RKI 2020d and Germany's age structure |
|  |  | 55-60: 0.998 |  |
|  |  | 60-65: 0.996 |  |
|  |  | 65-70: 0.983 |  |
|  |  | 70-75: 0.967 |  |
|  |  | 75-80: 0.936 |  |
|  |  | 80-85: 0.893 |  |
|  |  | 85-90: 0.829 |  |
|  |  | 90+: 0.701 |  |
|  |  | **total: 0.984** |  |
| Survival chance once at ICU | *surv_icu* | 0.7<br>(applied to all age groups) | Own calculations based on RKI 2020b; Nachtigall et al. 2020 |

## Appendix 2: Additional figures

**Figure A2:**
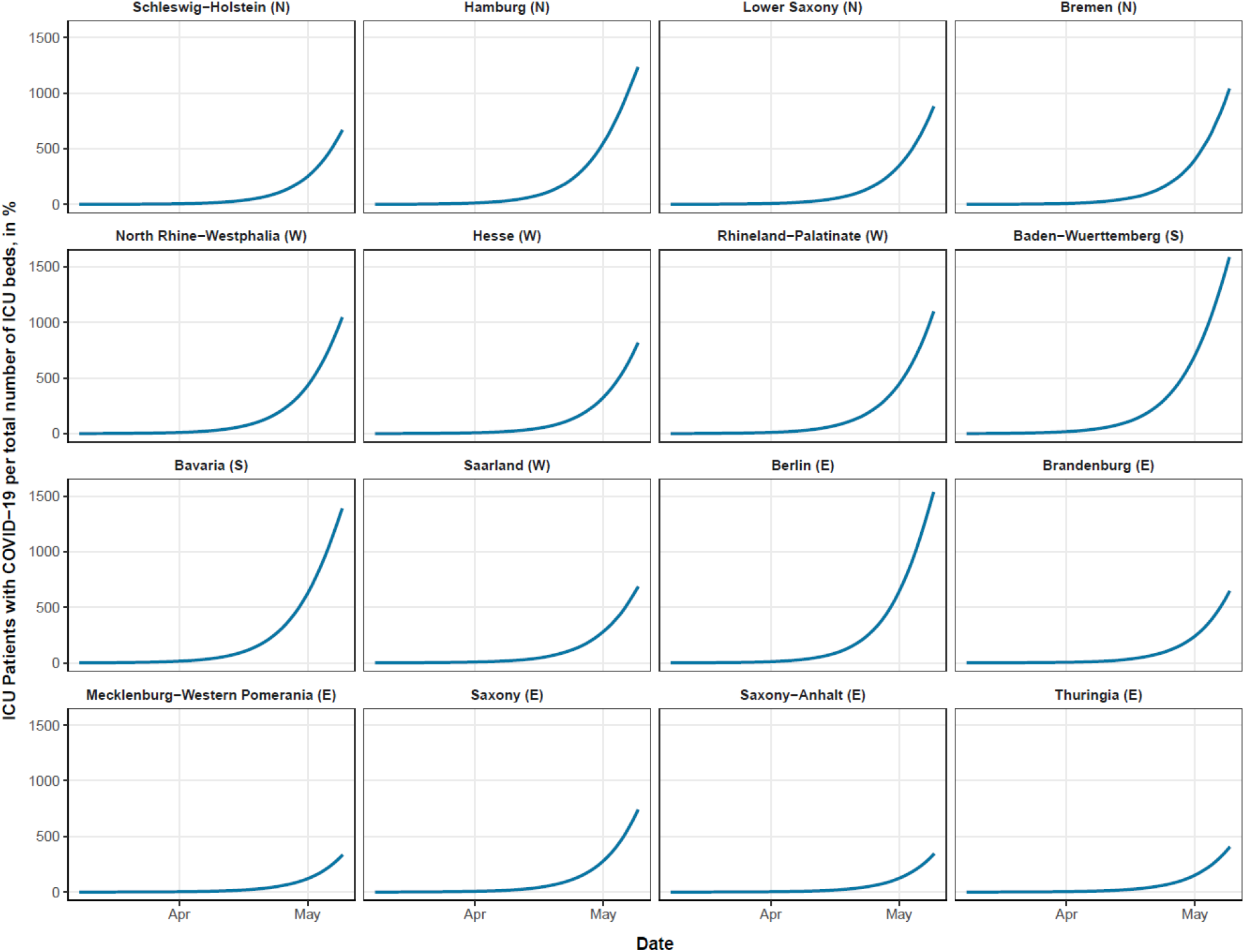
Forecast of COVID-19-related ICU demand in the uncontained setting (case 1), federal state level.

**Figure A3:**
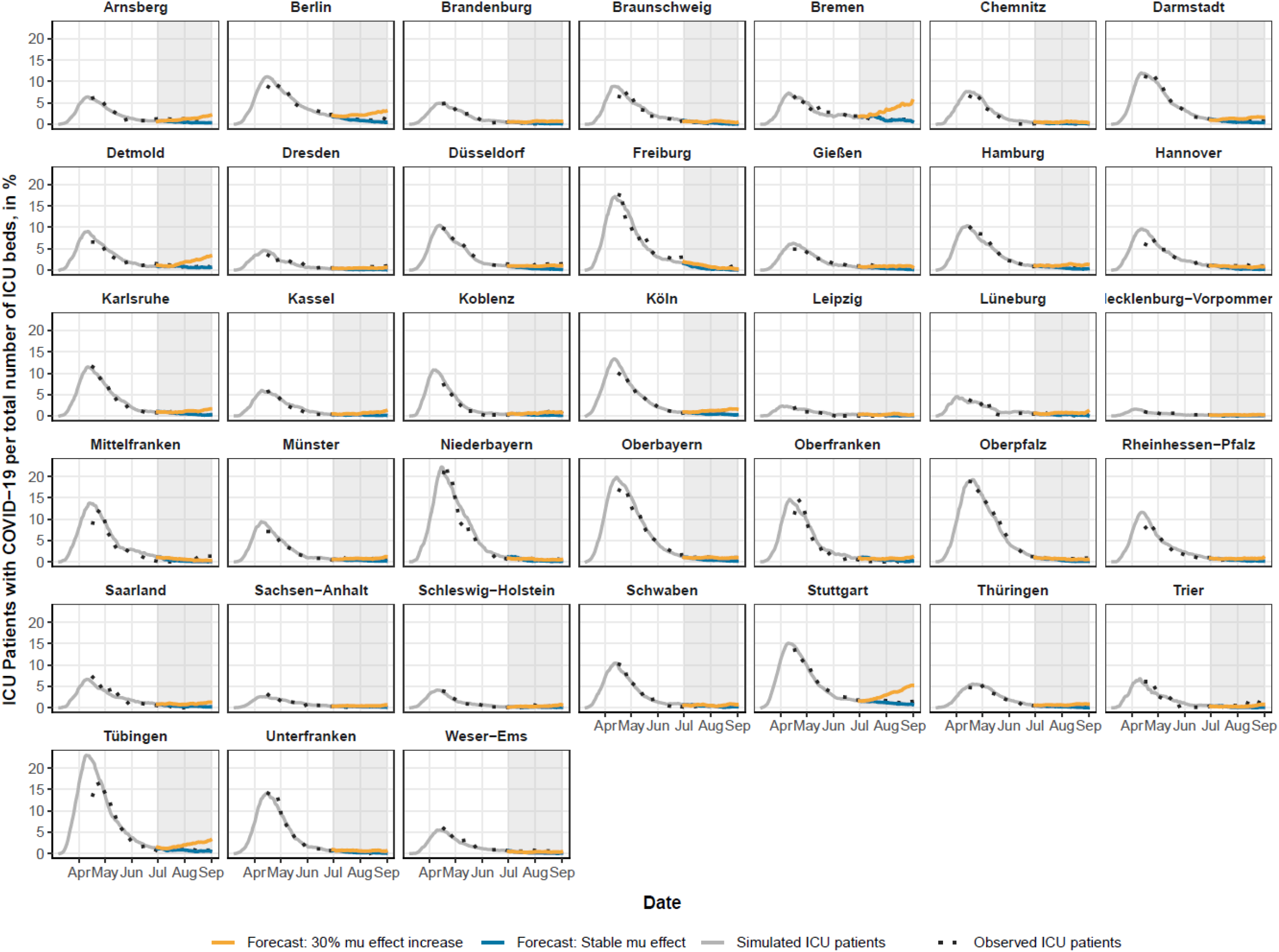
Forecast of COVID-19-related ICU demand in low-dynamic setting (case 2), NUTS-2 level.

**Figure A4:**
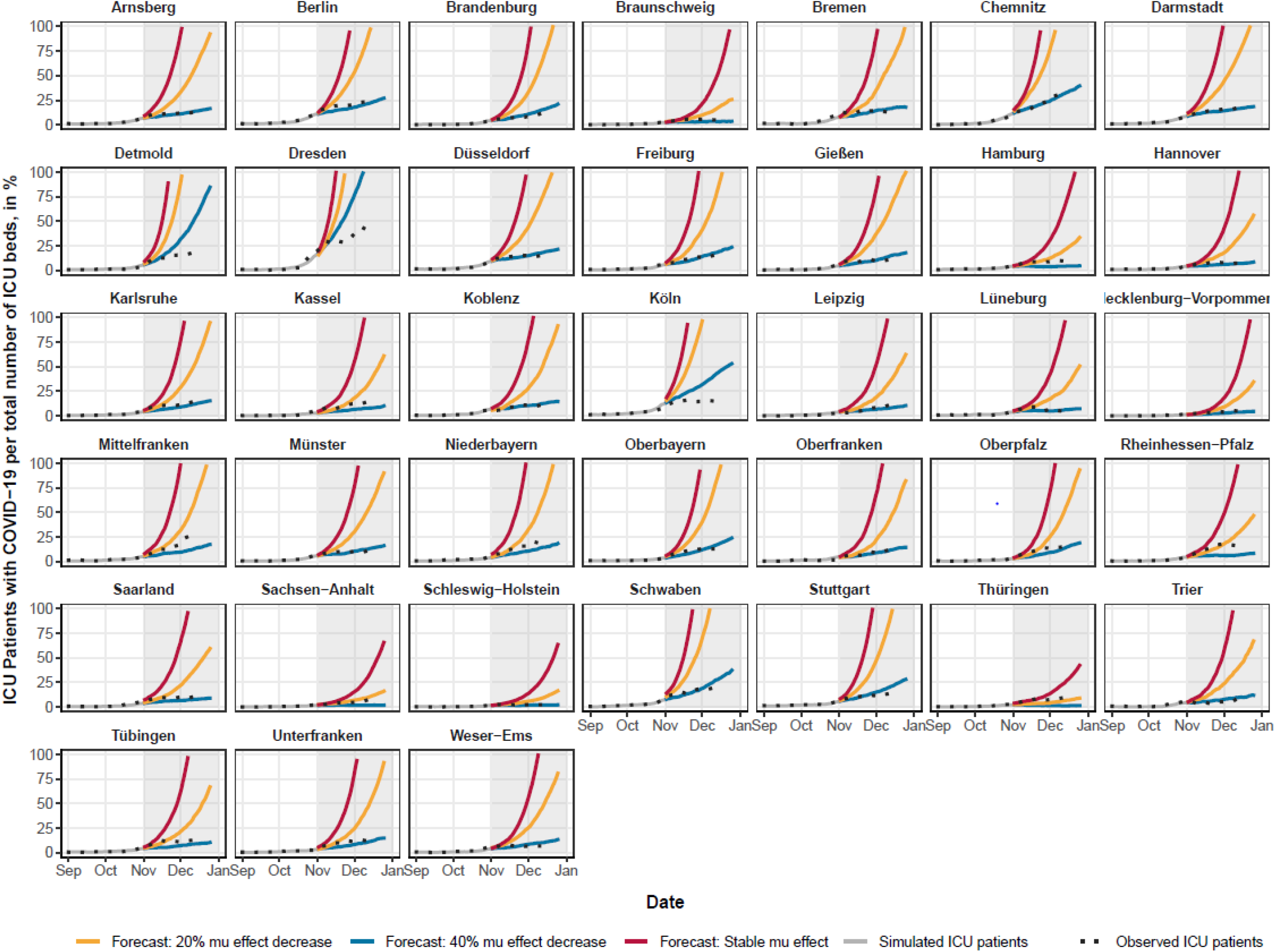
Forecast of COVID-19-related ICU demand in high-dynamic setting (case 3), NUTS-2 level.

**Figure A5:**
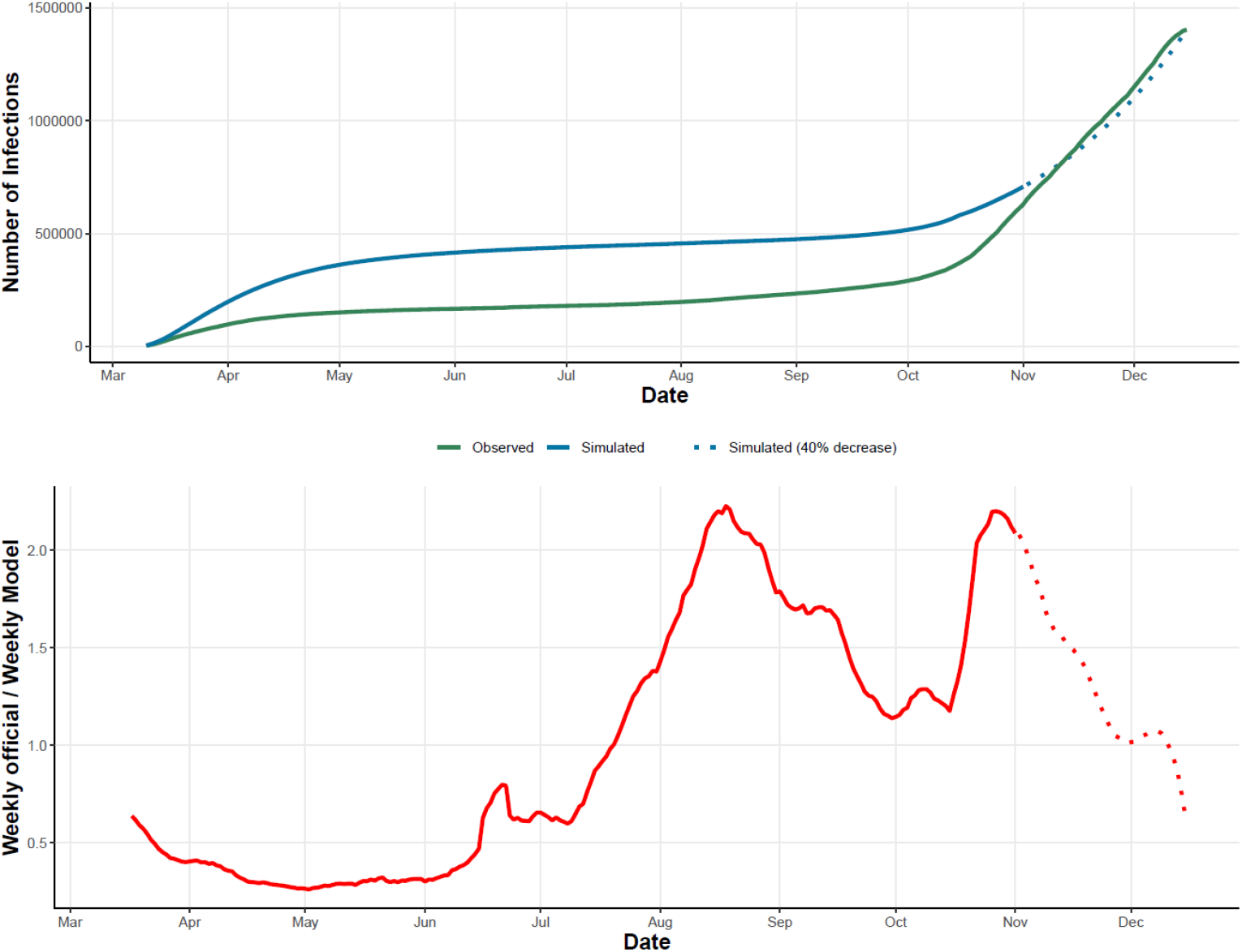
Number of observed and simulated infections, and their weekly share. Note: As our model is calibrated against ICU data, we are only able to determine the ICU-relevant infections levels. If, e.g., local outbreaks are concentrated among young persons, this would have only limited impact on ICU numbers, and would thus rather go undetected by our model. Nevertheless, it is interesting to see how the infections generated by our model differ from the registered number of infections. The registered number of infections were in the first wave in spring 2020 below 50% of the number of infections generated by our model. During the summer months, however, our model generated fewer infections than were actually reported, which is mainly attributed to a high share of young individuals being infected who are usually not send to ICUs.

**Figure A6:**
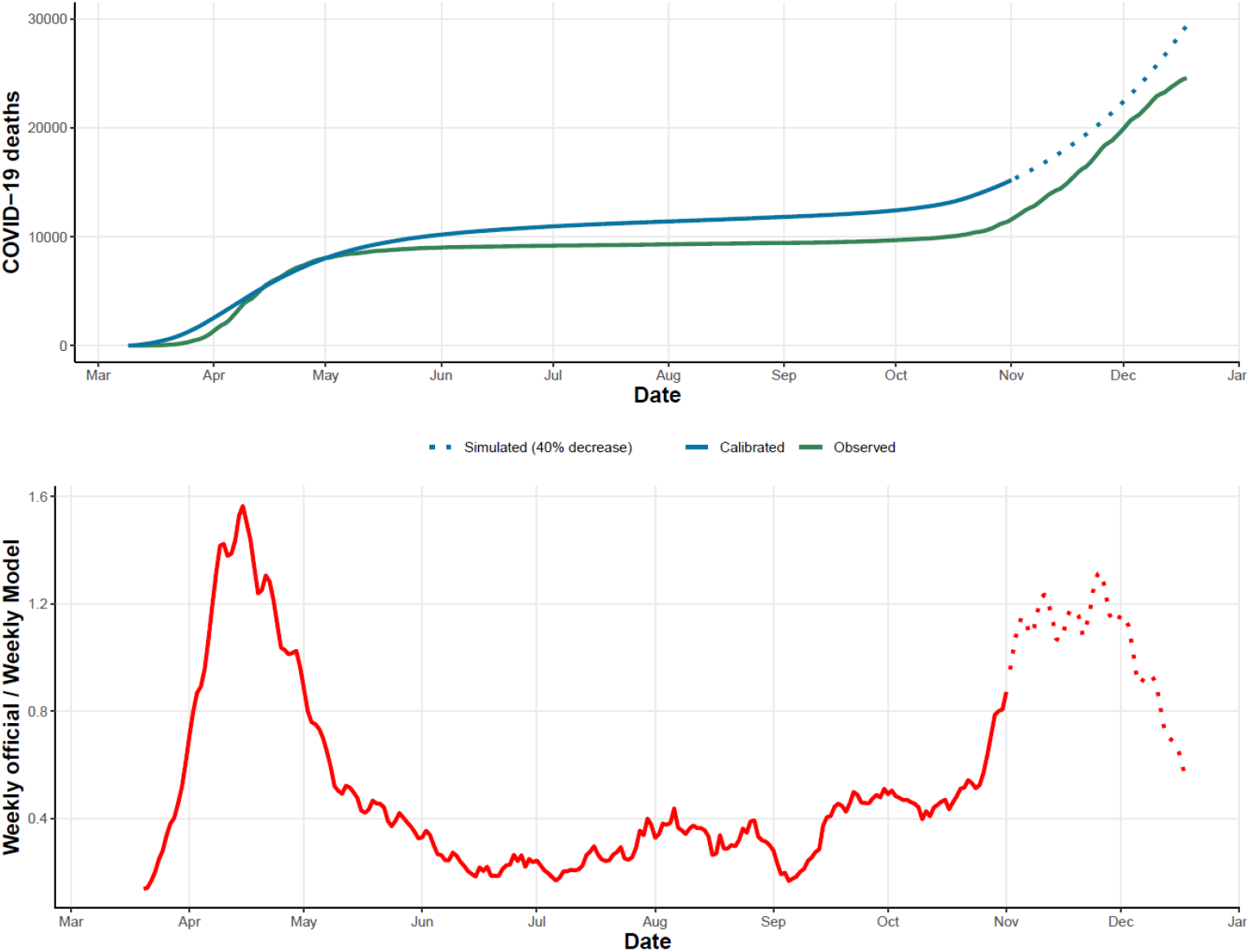
Number of observed and simulated deaths, and their weekly share.

**Figure A7:**
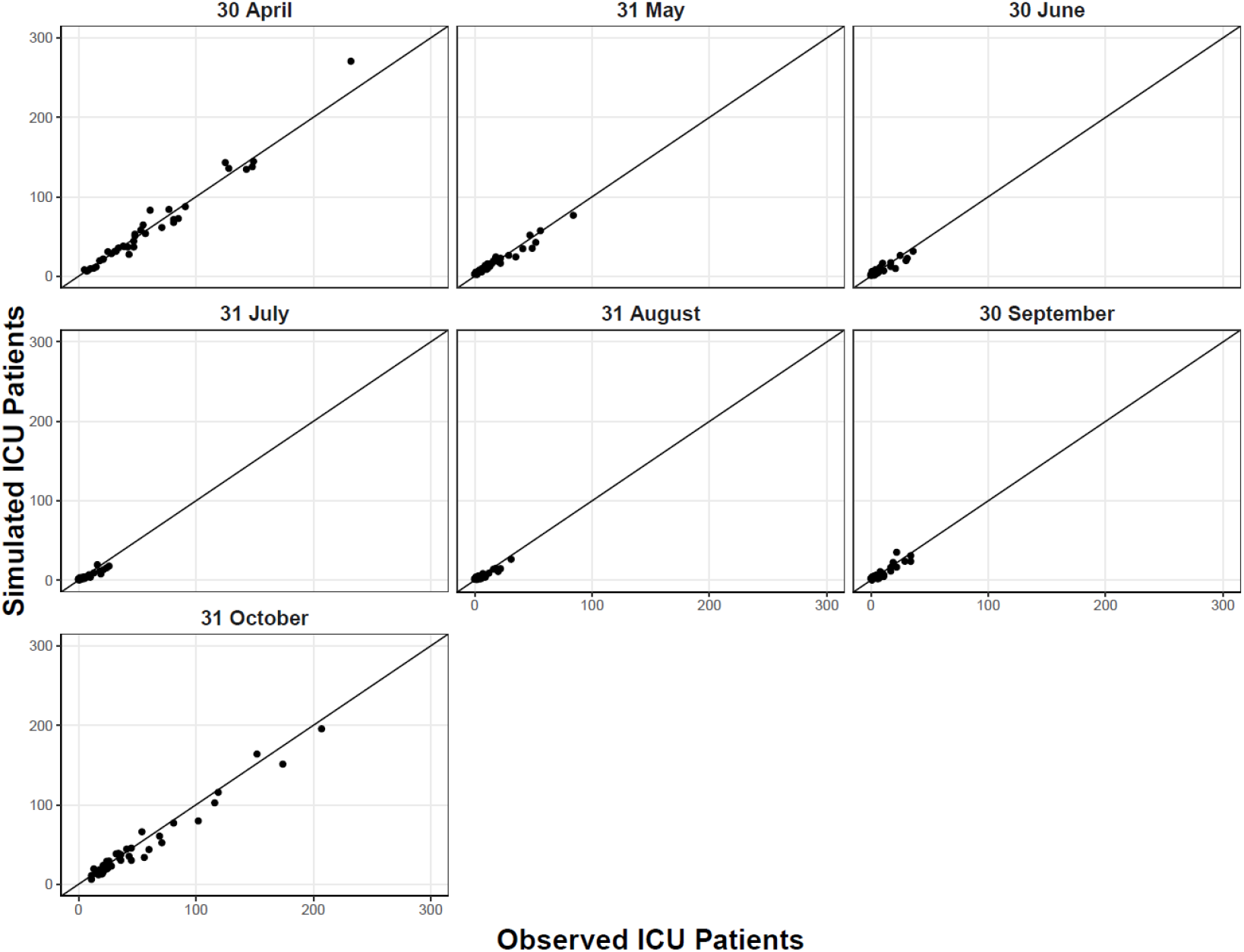
Relationship between observed and simulated ICU patients for different time points, NUTS-2 level.

**Figure A8:**
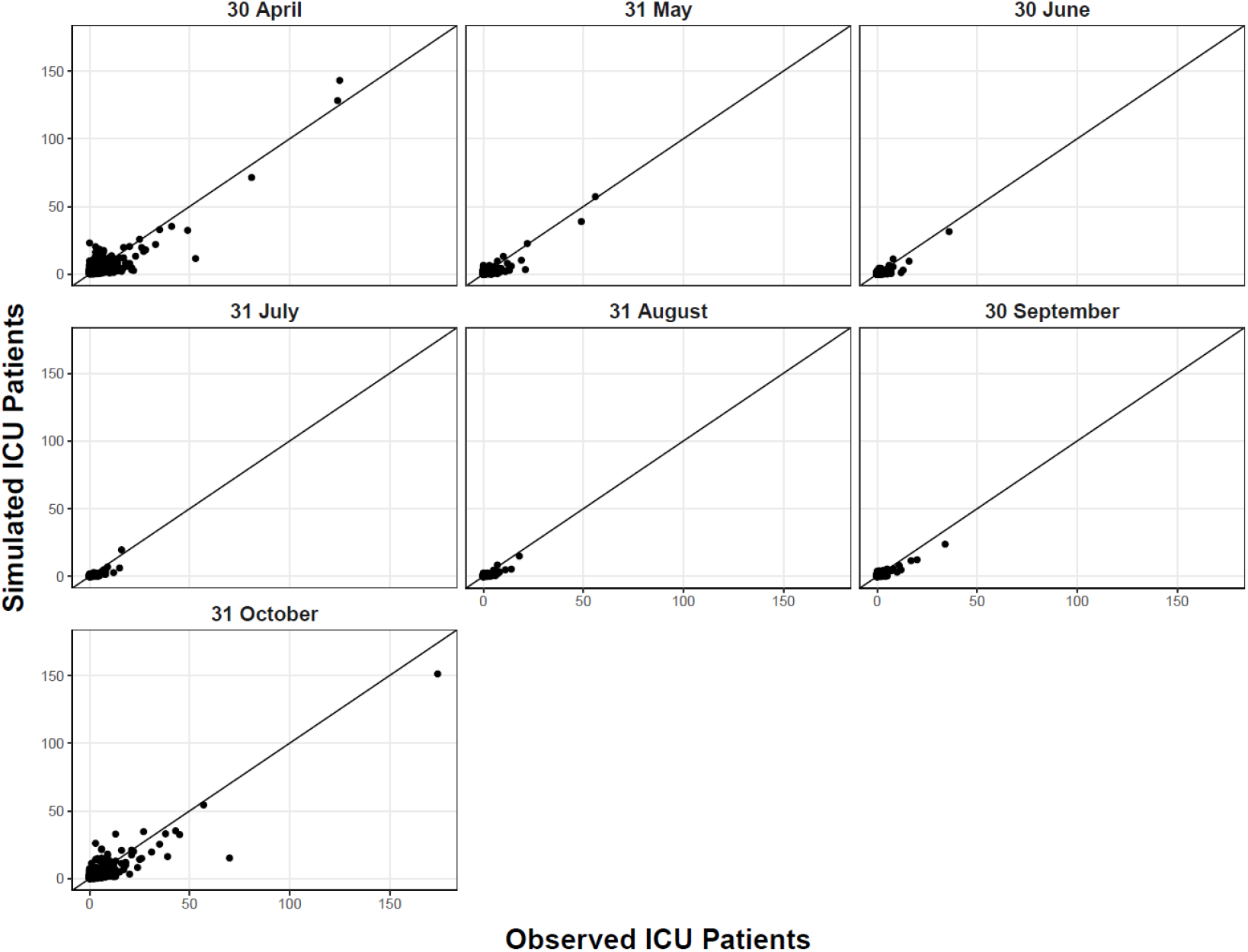
Relationship between observed and simulated ICU patients for different time points, district level.

**Figure A9:**
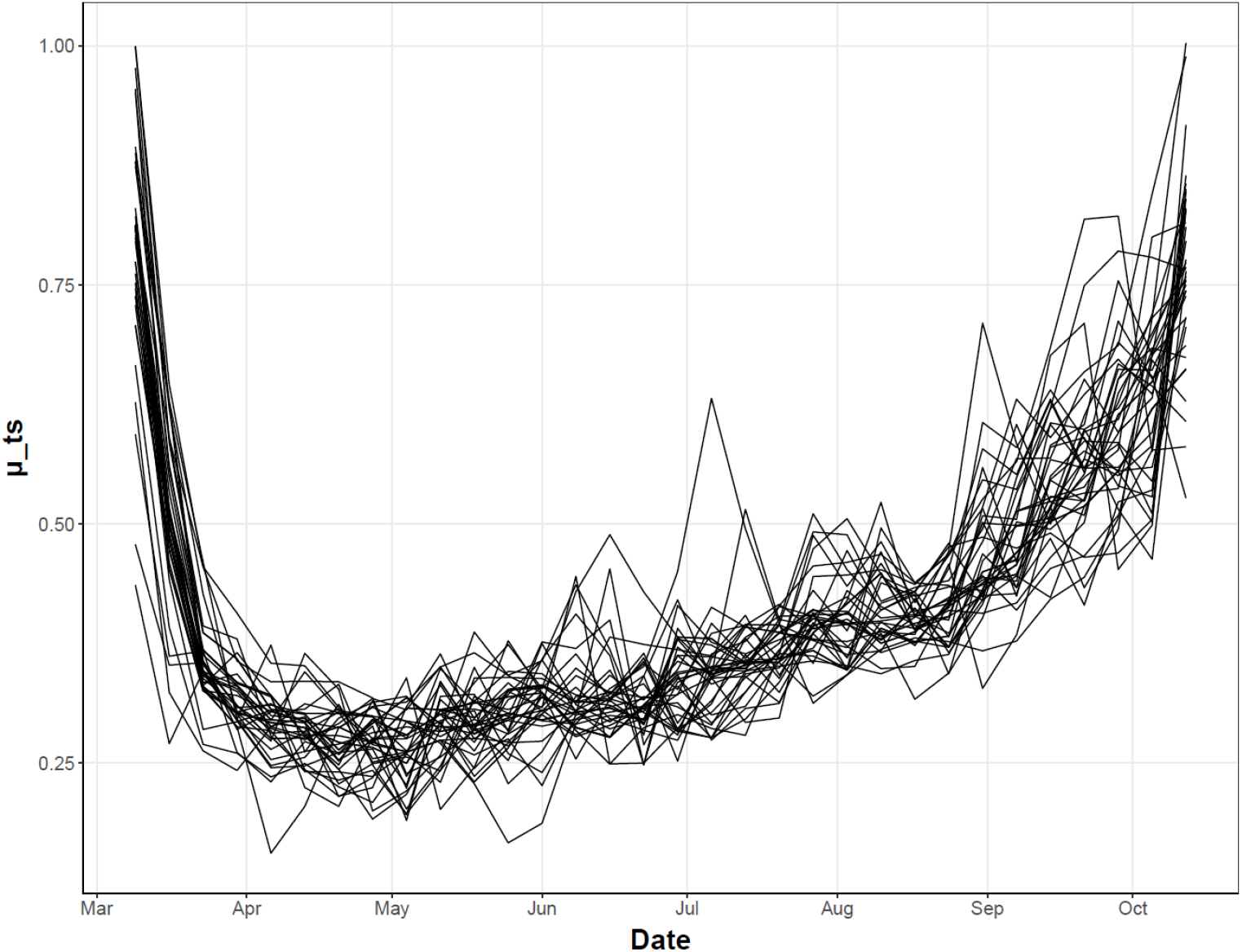
Development of *μ*_*ts*_ effects over time for the 38 NUTS-2 regions.

**Figure A10:**
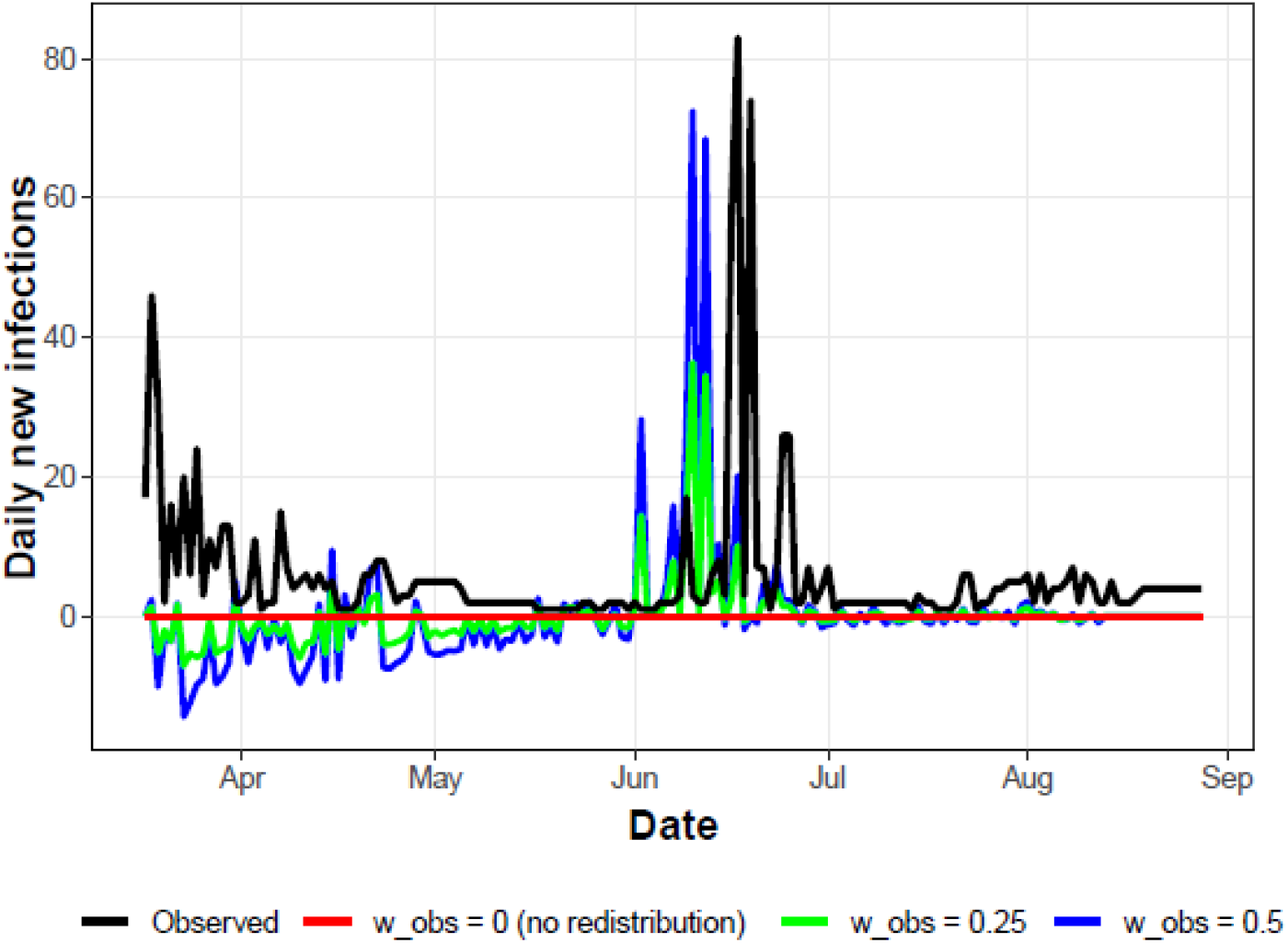
Example how *w_obs* redistributes newly generated infections within the model on the district level. Note: The plot shows the daily new infections in the district of Warendorf in western Germany which was affected by COVID-19 outbreak in a big slaughterhouse in June 2020 (see black line). Assigning the parameter *w_obs* a value of 0.5 (blue line), allows us to account for such local outbreaks by redistributing newly generated infections across Germany. The spike in the blue line occurs roughly five days earlier because we correct the RKI data for the beginning of the infection (see also Appendix 1).

## Literature

Alban, A., Chick, S. E., Dongelmans, D. A., Vlaar, A., Sent, D., & Study Group (2020). ICU capacity management during the COVID-19 pandemic using a process simulation. Intensive care medicine, 46(8), 1624–1626. https://doi.org/10.1007/s00134-020-06066-7

an der Heiden, M. & Buchholz, U. (2020). Modellierung von Beispielszenarien der SARS-CoV-2- Epidemie 2020 in Deutschland. 10.25646/6571.2

an der Heiden, M. & Hamouda, O. (2020). Schätzung der aktuellen Entwicklung der SARS-CoV-2- Epidemie in Deutschland – Nowcasting. Epidemiologisches Bulletin, 17, 10–15. 10.25646/669

Bauer, A. & Weber, E. (2020). COVID-19: how much unemployment was caused by the shutdown in Germany? Applied Economics Letters. https://doi.org/10.1080/13504851.2020.1789544

Bäuerle, A. et al. (2020). Increased generalized anxiety, depression and distress during the COVID-19 pandemic: a cross-sectional study in Germany. Journal of Public Health, 42(4), 672–678. https://doi.org/10.1093/pubmed/fdaa106

Bohk-Ewald, C., Dudel C., & Myrskylä, M. (2020). A demographic scaling model for estimating the total number of COVID-19 infections. arXiv. https://arxiv.org/abs/2004.12836

Brauer, F. (2017). Mathematical epidemiology: Past, present, and future. Infectious Disease Modelling. 2(2), 113–127. https://doi.org/10.1016/j.idm.2017.02.001

Carcione, J., Santos, J., Bagaini, C. & Ba, J. (2020). A simulation of a COVID-19 epidemic based on a deterministic SEIR model. arXiv. 2004.03575

Chin, V. et al. (2020). A case study in model failure? COVID-19 daily deaths and ICU bed utilisation predictions in New York state. European Journal of Epidemiology, 35, 733–742. https://doi.org/10.1007/s10654-020-00669-6

Chowdhury, R. et al. (2020). Dynamic interventions to control COVID-19 pandemic: a multivariate prediction modelling study comparing 16 worldwide countries. European Journal of Epidemiology, 35, 389–399. https://doi.org/10.1007/s10654-020-00649-w

Davies, N. G. et al. (2020). Effects of non-pharmaceutical interventions on COVID-19 cases, deaths, and demand for hospital services in the UK: a modelling study. The Lancet Public Health, 5(7), e375–e385. https://doi.org/10.1016/S2468-2667(20)30133-X

Dehning, J. et al. (2020). Inferring change points in the spread of COVID-19 reveals the effectiveness of interventions. Science, 369(160), 1–9. 10.1126/science.abb9789

Del Fava, E. et al. (2020). The differential impact of physical distancing strategies on social contacts relevant for the spread of COVID-19. medRxiv. https://doi.org/10.1101/2020.05.15.20102657

Dowd, J. et al. (2020). Demographic science aids in understanding the spread and fatality rates of COVID-19. PNAS, 117, 9696–9698. https://doi.org/10.1073/pnas.2004911117

Dudel, C. et al. (2020). Monitoring trends and differences in COVID-19 case fatality rates using decomposition methods: Contributions of age structure and age-specific fatality. PLoS ONE, 15(9): e0238904. https://doi.org/10.1371/journal.pone.0238904

Ferguson, N. et al. (2020). Report 9: Impact of non-pharmaceutical interventions (NPIs) to reduce COVID-19 mortality and healthcare demand. https://www.imperial.ac.uk/mrc-global-infectious-disease-analysis/covid-19/report-9-impact-of-npis-on-covid-19/

Flaxman, S. et al. (2020). Estimating the effects of non-pharmaceutical interventions on COVID-19 in Europe. Nature, 584, 257–261. https://doi.org/10.1038/s41586-020-2405-7

Fuchs-Schündeln, N., Kuhn, M., & Tertilt, M. (2020). The short-run macro implications of school and child-care closures (No. 13353). Institute of Labor Economics (IZA).

Ganyani, T. et al. (2020). Estimating the generation interval for coronavirus disease (COVID-19) based on symptom onset data, March 2020. Euro Surveill., 25(17), 2000257. https://doi.org/10.2807/1560-7917.ES.2020.25.17.2000257

Han, E. et al. (2020). Lessons learnt from easing COVID-19 restrictions: an analysis of countries and regions in Asia Pacific and Europe. The Lancet, 396(10261), 1525–1534. https://doi.org/10.1016/S0140-6736(20)32007-9

Haug, N. et al. (2020). Ranking the effectiveness of worldwide COVID-19 government interventions. medRxiv. https://doi.org/10.1101/2020.07.06.20147199

He, X. et al. (2020). Temporal dynamics in viral shedding and transmissibility of COVID-19. Nature Medicine, 26, 672–675. https://doi.org/10.1038/s41591-020-0869-5

Holmdahl, I. & Buckee, C. (2020). Wrong but Useful — What Covid-19 Epidemiologic Models Can and Cannot Tell Us. New England Journal of Medicine, 383, 303–305. https://doi.org/10.1056/NEJMp2016822

Ioannidis, J., Axfors, C. & Contopoulos-Ioannidis, D. (2020). Population-level COVID-19 mortality risk for non-elderly individuals overall and for non-elderly individuals without underlying diseases in pandemic epicenters. Environmental Research, 188. https://doi.org/10.1016/j.envres.2020.109890

Ioannidis, J., Cripps, S. & Tanner M. (2020). Forecasting for COVID-19 has failed. https://forecasters.org/blog/2020/06/14/forecasting-for-covid-19-has-failed/

Jewell N.P., Lewnard J.A., & Jewell B.L. (2020). Predictive Mathematical Models of the COVID-19 Pandemic: Underlying Principles and Value of Projections. JAMA, 323(19), 1893–1894. 10.1001/jama.2020.6585

IHME COVID-19 Forecasting Team, Murray, C.J.L. (2020). Forecasting COVID-19 impact on hospital bed-days, ICU-days, ventilator-days and deaths by US state in the next 4 months. medRxiv. https://doi.org/10.1101/2020.03.27.20043752

IHME COVID-19 Forecasting Team (2020). Modeling COVID-19 scenarios for the United States. Nat Med. https://doi.org/10.1038/s41591-020-1132-9

Kapitsinis, N. (2020). The underlying factors of the COVID-19 spatially uneven spread. Initial evidence from regions in nine EU countries. Regional Science Policy and Practice. 1–19. https://doi.org/10.1111/rsp3.12340

Karagiannidis, C. et al. (2020). Case characteristics, resource use, and outcomes of 10 021 patients with COVID-19 admitted to 920 German hospitals: an observational study. Lancet Respir Med, 8, 853–862. https://doi.org/10.1016/S2213-2600(20)30316-7

Kucharski, A. et al. (2020). Early dynamics of transmission and control of COVID-19: a mathematical modelling study. The Lancet Infectious Diseases, 20(5), 553–558. https://doi.org/10.1016/S1473-3099(20)30144-4

Levin, A. et al. (2020). Assessing the Age Specificity of Infection Fatality Rates for COVID-19: Systematic Review, Meta-Analysis, and Public Policy Implications. medRxiv. https://doi.org/10.1101/2020.07.23.20160895

Li, Y. et al. (2020a). The temporal association of introducing and lifting non-pharmaceutical interventions with the time-varying reproduction number (R) of SARS-CoV-2: a modelling study across 131 countries. The Lancet Infectious Diseases. https://doi.org/10.1016/S1473-3099(20)30785-4

Li, R. et al. (2020b). Substantial undocumented infection facilitates the rapid dissemination of novel coronavirus (SARS-CoV-2). Science, 368, 489–493. 10.1126/science.abb3221

McCabe, R. et al. (2020). Modelling ICU capacity under different epidemiological scenarios of the COVID-19 pandemic in three western European countries. Imperial College London (16-11-2020). https://doi.org/10.25561/84003.

Mense, A., & Michelsen, C (2020). Räumliche Ausbreitung von COVID-19 durch interregionale Verflechtungen [Spatial Interregional Spread of COVID-19 Through Commuter Interdependence]. Wirtschaftsdienst, 100(6), 416–421. https://doi.org/10.1007/s10273-020-2674-7

Moghadas, S. et al. (2020). The implications of silent transmission for the control of COVID-19 outbreaks. PNAS, 117(30), 17513–17515. https://doi.org/10.1073/pnas.2008373117

Nachtigall, I. et al. (2020). Clinical course and factors associated with outcomes among 1904 patients hospitalized with COVID-19 in Germany: an observational study. Clinical Microbiology and Infection, 26(12), 1663–1669. https://doi.org/10.1016/j.cmi.2020.08.011

Nadler, P. et al. (2020). An epidemiological modelling approach for COVID-19 via data assimilation. European Journal of Epidemiology, 35, 749–761. https://doi.org/10.1007/s10654-020-00676-7

Naumann, E. et al. (2020). COVID-19 policies in Germany and their social, political, and psychological consequences. European Policy Analysis. https://doi.org/10.1002/epa2.1091

Nepomuceno, M. et al. (2020). Besides population age structure, health and other demographic factors can contribute to understanding the COVID-19 burden. PNAS, 117, 13881–13883. https://doi.org/10.1073/pnas.2008760117

Perrotta et al. (2020). Behaviours and attitudes in response to the COVID-19 pandemic: Insights from a cross-national Facebook survey. medRxiv. https://doi.org/10.1101/2020.05.09.20096388

Prem, K. et al. (2020). The effect of control strategies to reduce social mixing on outcomes of the COVID-19 epidemic in Wuhan, China: a modelling study. The Lancet Public Health, 5(5), 261–270. https://doi.org/10.1016/S2468-2667(20)30073-6

Rhodes et al. (2012). The variability of critical care bed numbers in Europe. Intensive Care Medicine. 38, 1647–1653. https://doi.org/10.1007/s00134-012-2627-8

Robert Koch-Institute (2020a). Tagesdaten-CSV aus dem DIVI-Intensivregister. http://dx.doi.org/10.25646/7699

Robert Koch-Institute (2020b). Aktuelle DIVI-Intensivregisterdaten der letzten 24h (restricted access).

Robert Koch-Institute (2020c). SARS-CoV-2 Steckbrief zur Coronavirus-Krankheit-2019 (COVID-19). https://www.rki.de/DE/Content/InfAZ/N/Neuartiges_Coronavirus/Steckbrief.html

Robert Koch-Institute (2020d). COVID-19 Dashboard. https://npgeo-corona-npgeo-de.hub.arcgis.com/datasets/dd4580c810204019a7b8eb3e0b329dd6_0

Rodriguez-Morales, A. et al. (2020). Clinical, laboratory and imaging features of COVID-19: A systematic review and meta-analysis. Travel Medicine and Infectious Disease, 34, 1–13. https://doi.org/10.1016/j.tmaid.2020.101623

Römmele C. et al. (2020). Bettenkapazitätssteuerung in Zeiten der COVID-19-Pandemie : Eine simulationsbasierte Prognose der Normal- und Intensivstationsbetten anhand der deskriptiven Daten des Universitätsklinikums Augsburg [Bed capacity management in times of the COVID-19 pandemic : A simulation-based prognosis of normal and intensive care beds using the descriptive data of the University Hospital Augsburg]. Anaesthesist, 69(10), 717–725. https://doi.org/10.1007/s00101-020-00830-6

Ruktanonchai, N. W. et al. (2020). Assessing the impact of coordinated COVID-19 exit strategies across Europe. Science, 369(6510), 1465–1470. 10.1126/science.abc5096

Schlosser, F. et al. (2020). COVID-19 lockdown induces disease-mitigating structural changes in mobility networks. PNAS. https://doi.org/10.1073/pnas.2012326117

Scrucca, L. (2013). GA: A Package for Genetic Algorithms in R. Journal of Statistical Software, 53(4), 1–37. http://www.jstatsoft.org/v53/i04/

Stang, A. et al. (2020a). Geschätzte Nutzung von Intensivbetten aufgrund von COVID-19 in Deutschland im zeitlichen Verlauf. Deutsches Ärzteblatt, 117(19), 329–335. 10.3238/arztebl.2020.0329

Stang, A. et al. (2020b). Excess mortality due to COVID-19 in Germany. Journal of Infection, 81, 797–801. https://doi.org/10.1016/j.jinf.2020.09.012

Streeck, H. et al. (2020). Infection fatality rate of SARS-CoV2 in a super-spreading event in Germany. Nature Communications. https://doi.org/10.1038/s41467-020-19509-y

Teslya, A., Pham, T. M., Godijk, N. G., Kretzschmar, M. E., Bootsma, M., & Rozhnova, G. (2020). Impact of self-imposed prevention measures and short-term government-imposed social distancing on mitigating and delaying a COVID-19 epidemic: A modelling study. PLoS medicine, 17(12), e1003499. https://doi.org/10.1371/journal.pmed.1003166

Thomas, L. et al. (2020). Spatial heterogeneity can lead to substantial local variations in COVID-19 timing and severity. PNAS, 117(39), 24180–24187. https://doi.org/10.1073/pnas.201165611

Tizzoni, M. et al. (2014). On the use of human mobility proxies for modeling epidemics. PLoS Comput Biol, 10(7), e1003716. https://doi.org/10.1371/journal.pcbi.1003716

Wu, Z. & McGoogan, J. (2020). Characteristics of and important lessons from the Coronavirus disease 2019 (COVID-19) Outbreak in China summary of a report of 72 314 cases from the Chinese Center for Disease Control and Prevention. JAMA, 323 (13), 1239–1242. https://doi.org/10.1001/jama.2020.2648

